# Population pharmacokinetics and exposure-response analysis of sotrovimab in the early treatment of COVID-19

**DOI:** 10.1101/2022.11.23.22282478

**Authors:** Jennifer E. Sager, Asma El-Zailik, Julie Passarell, Stefan Roepcke, Xiaobin Li, Melissa Aldinger, Ahmed Nader, Andrew Skingsley, Elizabeth L. Alexander, Wendy W. Yeh, Erik Mogalian, Chad Garner, Amanda Peppercorn, Adrienne E. Shapiro, Maribel Reyes

**Affiliations:** Vir Biotechnology, Inc., San Francisco, CA, USA; Cognigen Division, Simulations Plus, Inc., Buffalo, NY, USA; GSK, Upper Providence, PA, USA; GSK, Brentford, UK; GSK, Cambridge, Massachusetts, USA; Fred Hutchinson Cancer Center, Seattle, WA, USA

**Keywords:** Exposure response, infectious disease, monoclonal antibodies, population pharmacokinetics

## Abstract

Sotrovimab is a recombinant human monoclonal antibody that has been shown to prevent progression to hospitalization or death from severe disease in non-hospitalized high-risk patients with mild-to-moderate COVID-19 following either intravenous (IV) or intramuscular (IM) administration. Population pharmacokinetic (popPK) and exposure-response (ER) analyses were performed to characterize sotrovimab PK and the relationship between exposure and response (probability of progression), as well as covariates that may contribute to between-participant variability in sotrovimab PK and efficacy following IV or IM administration. Sotrovimab PK was described by a two-compartment model with linear elimination; IM absorption was characterized by a sigmoid absorption model. PopPK covariate analysis led to the addition of the effect of body weight on systemic clearance and peripheral volume of distribution, sex on IM bioavailability and first-order absorption rate (KA), and body mass index on KA. However, the magnitude of covariate effect was not pronounced and was therefore not expected to be clinically relevant based on available data to date. For ER analysis, sotrovimab exposure measures were predicted using the final popPK model. An ER model was developed using the exposure measure of sotrovimab concentration at 168 hours that described the relationship between exposure and probability of progression within the ER dataset for COMET-TAIL. The number of risk factors (≤1 vs >1) was incorporated as an additive shift on the model-estimated placebo response but had no impact on overall drug response. Limitations in the ER model may prevent generalization of these results to describe the sotrovimab exposure-progression relationship across SARS-COV-2 variants.

## INTRODUCTION

Preventing infection and reducing the risk of progression of coronavirus disease 2019 (COVID-19) remains an urgent public health priority. While currently available vaccines have been shown to be efficacious in preventing severe COVID-19,^1, 2^ certain subgroups of individuals remain at higher risk of severe COVID-19, resulting in hospitalization and increased risk of mortality. These subgroups include people ≥55 years of age, those with unvaccinated status, and those with comorbidities, including cardiovascular disease, diabetes, renal disease, neurological conditions, and immune suppression.^3, 4, 5, 6^ Care of patients with severe COVID-19 is resource intensive and poses a substantial burden on hospital reserves; thus, preventing progression to severe disease among high-risk patients with mild or moderate COVID-19 is an important goal of treatment.^7, 8^

Sotrovimab was developed to treat mild to moderate COVID-19 in non-hospitalized patients at high risk of disease progression.^9, 10^ Sotrovimab is a recombinant human immunoglobulin G1 (IgG1) monoclonal antibody (mAb) that binds to a conserved epitope within the virus spike protein receptor binding domain of severe acute respiratory syndrome coronavirus-2 (SARS-CoV-2).^11, 12^ The fragment crystallizable domain of sotrovimab includes the two amino acid “LS” modification that has been shown to extend antibody half-life in studies of other mAbs and may enhance distribution of sotrovimab to the respiratory mucosa.^13, 14, 15^

The efficacy of early treatment with sotrovimab in preventing the progression of COVID-19 in those who are at risk for hospitalization or death has been demonstrated in two pivotal clinical trials of non-hospitalized patients with mild to moderate COVID-19 at high risk for disease progression.^9, 10, 16^ In the COMET-ICE study, a 500-mg dose of sotrovimab, administered intravenously (IV), demonstrated a relative risk reduction of 79% in hospitalization greater than 24 hours for acute management of any illness, or death due to any cause through Day 29 compared with placebo.^10^ In the COMET-TAIL clinical trial, efficacy of intramuscular (IM) sotrovimab 500 mg demonstrated non-inferiority^a^ compared with IV sotrovimab given at the same dose, with a low incidence of COVID-19 progression observed for both routes of administration.^16^ Sotrovimab was well tolerated in these studies, specifically with a low frequency of infusion-related reactions and only mild and transient injection-site reactions.

Understanding factors that influence both exposure and response of a given treatment option is important in informing clinical usage. Population pharmacokinetic (popPK) and exposure-response (ER) analyses were conducted to 1) characterize the popPK of sotrovimab following IV and IM administration, 2) identify and quantify the effects of intrinsic and extrinsic factors influencing the pharmacokinetics (PK) of sotrovimab using systematic covariate analysis, 3) describe the relationship between sotrovimab exposure and probability of progression of COVID-19 based on IV and IM data from the COMET-TAIL study, and 4) identify clinical covariates that influence variability in efficacy response.

## METHODS

### Clinical studies

Data for popPK analysis were derived from four clinical studies in non-hospitalized COVID-19 patients: COMET-ICE (ClinicalTrials.gov identifier, NCT045450060), COMET-TAIL (NCT04913675), COMET-PEAK (NCT04779879), and BLAZE-4 (NCT04634409), together with one study in healthy volunteers of Japanese or Caucasian descent (NCT04988152). Data for the sotrovimab ER analysis were derived from COMET-TAIL. Details of the clinical study designs are published or available elsewhere.^9, 10, 16, 17, 18, 19^

All participants who received sotrovimab and had at least one measurable concentration of drug were included in the popPK analysis dataset. Study participants in the analysis dataset received sotrovimab as single IV (500 mg) or IM (250 mg or 500 mg) doses. In the BLAZE-4 study, sotrovimab (500 mg IV) was administered in combination with bamlanivimab. All studies were conducted in accordance with the ethical principles derived from the Declaration of Helsinki and Council for International Organizations of Medical Sciences International Ethical Guidelines, applicable International Council for Harmonisation Good Clinical Practice guidelines, and applicable laws and regulations. Ethics approval was obtained from institutional review boards and ethics committees. Written informed consent was provided prior to study entry; participants less than 18 years of age signed an assent form, and a parent/guardian provided written consent.

### PK sampling and assay

Venous blood samples were obtained at different times prior to and after treatment administration (**Table S1**). Serum sotrovimab concentrations were determined using a validated electrochemiluminescent method validated on the Meso Scale Discovery (Rockville, MD, USA), with a lower limit of quantification of 0.1 μg/mL.

### PopPK modeling software and dataset

PopPK modeling of sotrovimab concentration-time data was performed using NONMEM (version 7.3). A pooled NONMEM-ready dataset was constructed using SAS (version 9.4 or higher). The dataset contained dosing history, infusion rate, sotrovimab plasma concentration data, relevant laboratory baseline values, and demographic and covariate information.

### PopPK model development

Model development was performed in a two-stage approach. An adequate model for sotrovimab PK was first developed with data following IV administration, after which data collected following IM administration was added to the analysis and the model was extended to include absorption processes specific to the IM route of administration. Initial development of the joint IV and IM popPK model included only participants with dense PK data. At the end of base model development, parameter estimation was performed on the entire dataset including all participants with dense or sparse data.

Exploratory analysis and prior knowledge of typical PK of therapeutic antibodies informed the selection of the functional form of the base structural model. Initially, a mammillary two-compartment model was assessed for appropriateness in describing the PK of sotrovimab. The variability model included random effect terms on the PK parameters elimination clearance (CL), central and peripheral volume of distribution (V2 and V3, respectively), absorption rate (KA), and bioavailability after IM injection (F_IM_) to describe the interindividual variability (IIV), and a combined additive and proportional error model to describe the residual variability (RV). Model evaluation was based on model diagnostics, goodness-of-fit (GOF) plots, and simulation-based visual predictive checks (VPCs). Various alternative models were applied to the data and assessed for their capacity to sufficiently characterize the popPK of sotrovimab, as needed.

Absolute estimates of disposition parameters following IV administration of sotrovimab were available from the clinical study data. Additionally, the rate and extent (ie, absolute bioavailability) of sotrovimab absorption after IM injections were estimated. A first-order model for sotrovimab absorption after IM administration was initially tested, followed by more complex absorption models.

Following the development of an appropriate base structural model, the influence of covariates on selected parameters was evaluated using a systematic forward inclusion and backward elimination approach. Covariates were added sequentially to the base model starting with the covariate contributing the most significant change in the minimum value of the objective function (VOF) (smallest *p* < 0.01) and a reduction in IIV in the parameter of interest of at least 5%. This process was repeated until there were no further covariates that produced significant changes in the VOF. Each covariate’s significance was tested individually with backward elimination until all remaining covariates were significant (change in VOF of at least 10.83; *p* < 0.001). Covariates investigated included age, sex, self-reported racial classification,^20^ disease state (healthy vs COVID-19), body weight, body mass index (BMI), hepatic function category (National Cancer Institute [NCI] classification), renal function category (based on estimated glomerular filtration rate; Modification of Diet in Renal Disease equation), serum albumin concentration, concomitant use of dexamethasone and/or remdesivir, baseline viral load, and sotrovimab clinical trial material (Gen 1 pool vs Gen 2 clonal cell line) (**Table S2**). In the healthy volunteer study, no serum albumin data were available, so covariate analysis for this factor was restricted to subjects from the remaining studies. For continuous variables, missing values were replaced by the respective study- and sex-specific median values in the dataset. For categorical variables, missing values were grouped with the respective unspecified category, eg, “unknown” or “other”.

The reduced multivariable model, including all significant covariates, was evaluated for any remaining biases in the IIV and RV error models. Adequacy of the final model was evaluated using a simulation-based VPC method. Utilizing NONMEM, the final model was used to simulate 500 replicates of the analysis dataset sufficient to achieve at least 100,000 patients per stratum of the VPC. Statistics of interest were calculated from simulated and observed data for comparison.

To characterize the impact of intrinsic and extrinsic factors on sotrovimab PK, simulations were performed based on the final popPK model and individual Bayesian estimates of PK parameters following actual treatment received. Numerical integration was performed to predict sotrovimab maximum concentration (C_max_), concentration at 96 hours (C_96h_), and concentration at 168 hours (C_168h_) for each patient. Summary statistics of the simulated exposures were calculated and stratified by covariate group (discrete covariates) or quantile (continuous covariates). Clinically meaningful impact on sotrovimab PK was determined based on geometric mean ratios (GMRs) and 90% confidence intervals (CIs) when compared to clinical bounds of 0.5 and 2.0 relative to the reference group.

### ER modeling

For the COMET-TAIL ER analysis, a NONMEM-ready dataset was constructed, which included sotrovimab dosing (dose, timing), treatment assignment, efficacy responses, demographic data, and clinical covariates. Sotrovimab exposure measures were predicted using the final popPK model and included predicted serum concentrations 24, 48, 72, 96, and 168 hours postdose, average concentrations and area under the curve (AUC) from time zero to 24, 48, 72, 96 and 168 hours after the dose, as well as AUC from time zero to Day 28 postdose (AUC_0-Day28_). Patients excluded from the popPK analysis were also excluded from the ER analysis. The endpoint used for the COMET-TAIL ER modeling included the primary efficacy endpoint from COMET-TAIL, which was the progression of COVID-19 through Day 29 as defined by hospitalization >24 hours for acute management of illness due to any cause or death.

The influence of covariates on the probability of progression of COVID-19 through Day 29 was evaluated using forward selection with α = 0.01. Covariates evaluated included age, sex, BMI, number of risk factors (inclusive of age and BMI), number of other risk factors (exclusive of age and BMI), symptom duration (continuous and categorical), baseline viral load, and route of administration (IV or IM).

Separate logistic regression models were developed for each exposure measure to determine if sotrovimab exposure was a statistically significant (α = 0.05) predictor of the probability of the progression endpoint. The exposure measure(s) selected for inclusion in the base logistic regression models was chosen based on statistical assessment in addition to clinical considerations.

Following forward selection, the full logistic regression model was used to predict the probability of the efficacy endpoint for various levels of categorical variables, or over the observed range of each continuous covariate that was statistically significant. This full model was evaluated for any remaining biases using a simulation-based VPC method.

## RESULTS

### Final popPK analysis dataset

A total of 1,984 participants contributed 14,269 sotrovimab concentration measurements to the popPK model. The final popPK dataset included 11,772 samples. A total of 2,497 samples were excluded from the analysis due to missing sample date and/or time information, duplicate sample date and/or time, predose and postdose below the lower limit of quantification samples, predose measurable concentration, or analyst-identified outliers (conditional weighted residuals [CWRES] <-5 or CWRES >5). PK samples were also excluded from the analysis if they were deemed nonphysiological or anomalous measured concentrations (<50 μg/mL or >500 μg/mL, assuming typical blood volumes [1 to 10 L]). These records were retained but flagged in the data file and excluded during the analysis. The numbers of participants and concentrations available by study and treatment group are presented in **Table S3**.

### PopPK demographics and baseline characteristics

Median age was 49 years, 44.9% were male, and 88.4% were White; median body weight was 83.6 kg, and median BMI was 30.4 kg/m^2^ (**Table S4**). The analysis included 1,891 patients with COVID-19 and 38 healthy volunteers. In total, 1,415 participants had normal renal function, while 401, 65, and 5 participants had mild, moderate, and severe renal impairment, respectively. Moreover, 1,393 participants had normal hepatic function, and 487 and 5 participants had mild and moderate hepatic impairment (NCI criteria), respectively.

### PopPK analysis

#### Base model

The PK of sotrovimab was best described by a two-compartment base model with first-order elimination. The absorption of the IM data was best described by a sigmoid absorption model, which was implemented using a zero-order input process into a depot compartment followed by first-order absorption into the central compartment (**Figure 1**).

**FIGURE 1.**
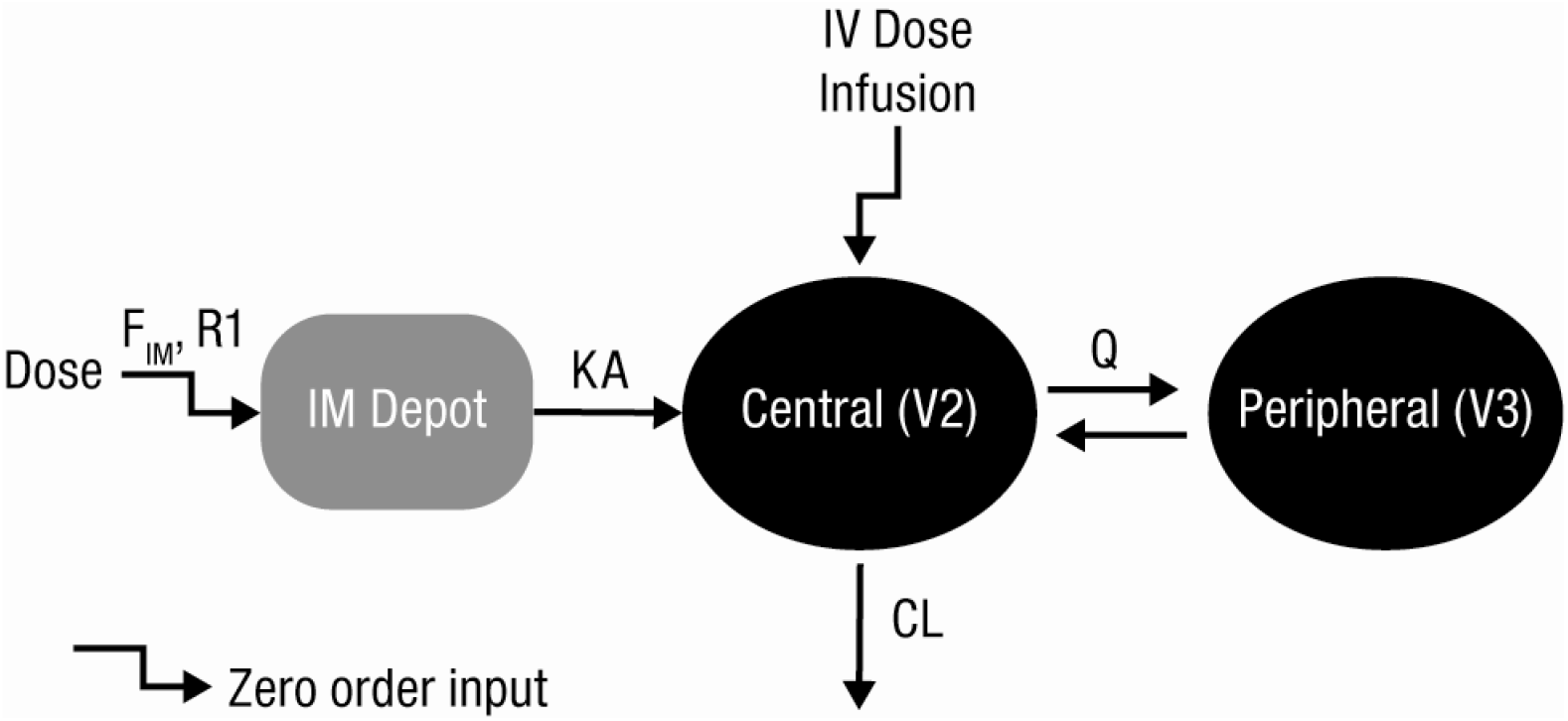
Base population PK model for IV and IM sotrovimab. CL, elimination clearance; F_IM_, bioavailability after IM injection; IM, intramuscular; IV, intravenous; KA, first-order rate of absorption of the intramuscular absorption compartment; PK, pharmacokinetic; Q, distribution clearance; R1, zero-order input rate for first IM absorption compartment; V2, central volume of distribution; V3, peripheral volume of distribution.

Various models were tested to characterize the IIV of the PK of sotrovimab, including IIV terms on V2, V3, CL, KA, and F_IM_. The constant coefficient of variation (CCV) and combined CCV and additive error models were tested to characterize the RV. IIV terms were added to CL, V2, V3, KA, and F_IM_ using the full covariance matrix. RV was described by a combined additive and CCV RV model. All parameters were estimated with good precision (percent relative standard error [%RSE] <8%), except the logit of F_IM_ (124 %RSE). Parameter shrinkage for the IIV parameters was 11.2% to 33.0%. The magnitude of the residual errors was moderate (%CV <14% for concentrations >10 μg/mL).

#### Final model

Covariate analysis via a stepwise forward selection and backward elimination approach led to the addition of the effect of body weight on systemic CL and V3, sex on F_IM_ and KA, and BMI on KA (**Table 1**). Variability in sotrovimab PK was additionally described by IIV on CL, V2, V3, F_IM_, and KA with a full covariance matrix and an additive plus CCV RV model. The final popPK parameter estimates, standard errors, and covariate effects are shown in **Table 1**. All fixed effect parameters were estimated with good precision (≤20% RSE). The magnitudes of the IIV were 29-57 %CV for CL, V2, V3 F_IM_, and KA. With a few exceptions, the random effect parameters were estimated with good precision (<28% RSE).

**TABLE 1.**
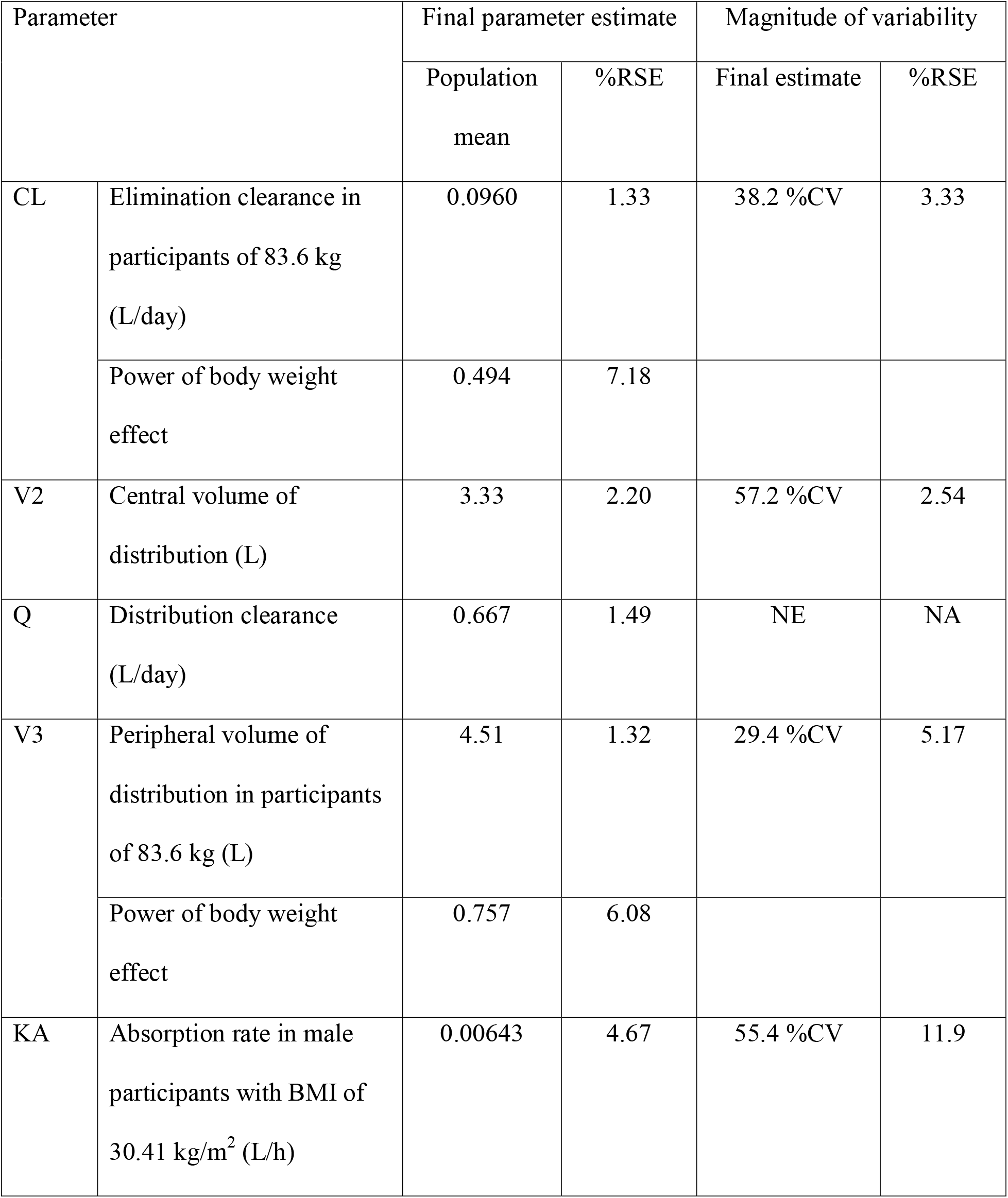
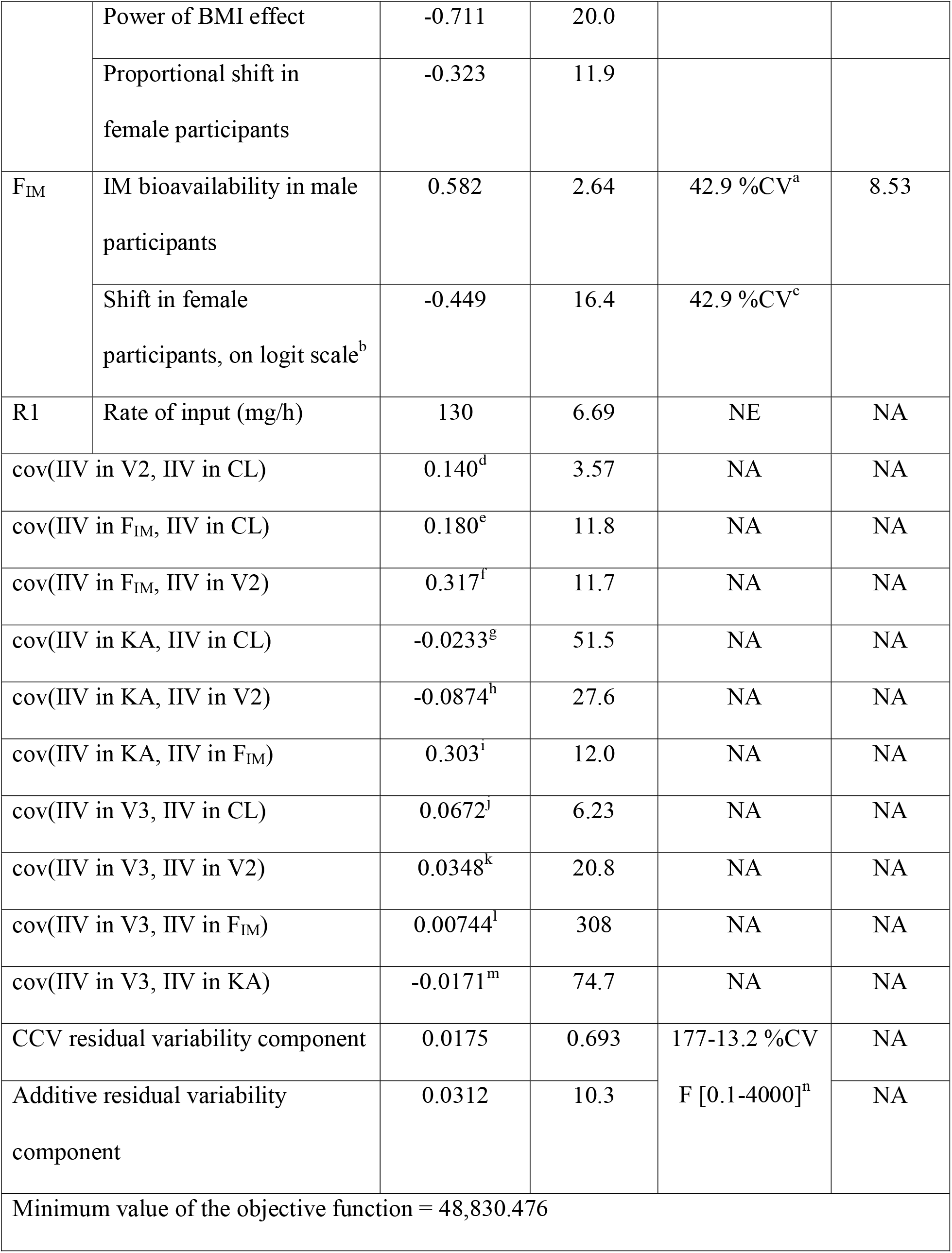
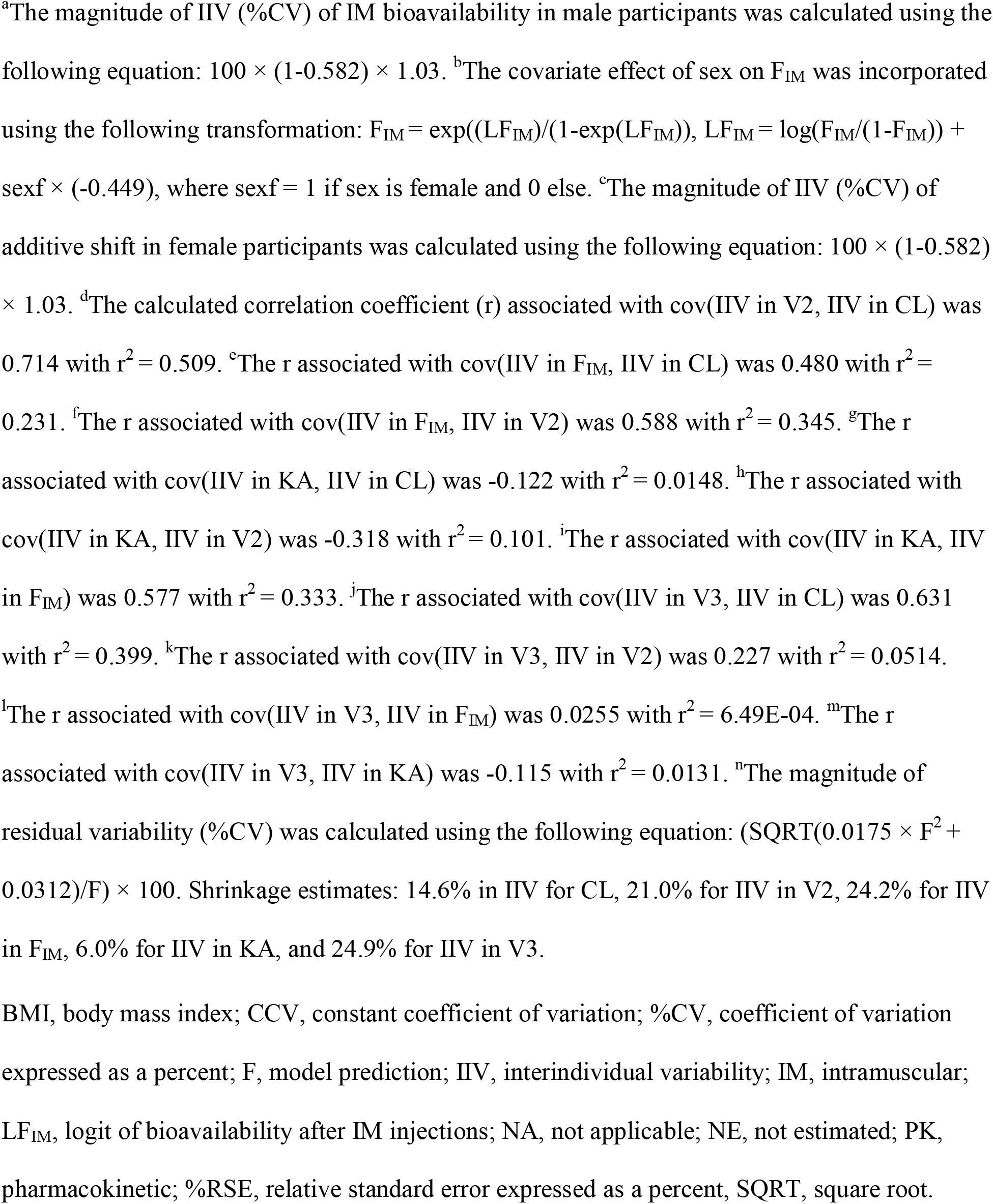
Final population PK parameter estimates and covariate effects

Based on the final model, the typical value for systemic CL was 0.096 L/day and V2 was 3.33 L. The rate of absorption following IM dosing was 0.00643 L/h. According to the model, the typical value of bioavailability in male participants was 0.582 and in female participants was 0.471. The model-estimated median half-life was 61.2 days (**Table S5**). GOF plots for the final popPK model, stratified by route of administration, show that the model adequately described the observed data for both routes of administration (**Figure S1**). VPC plots stratified by route and dose showed that the observed concentrations were mostly contained within the range of the 5^th^ and 95^th^ percentiles of the model-predicted concentration values (**Figure 2**). Overall, the final model captured the central tendency (median) and the extent of variability of the observed PK data following IV and IM routes of administration well.

**FIGURE 2.**
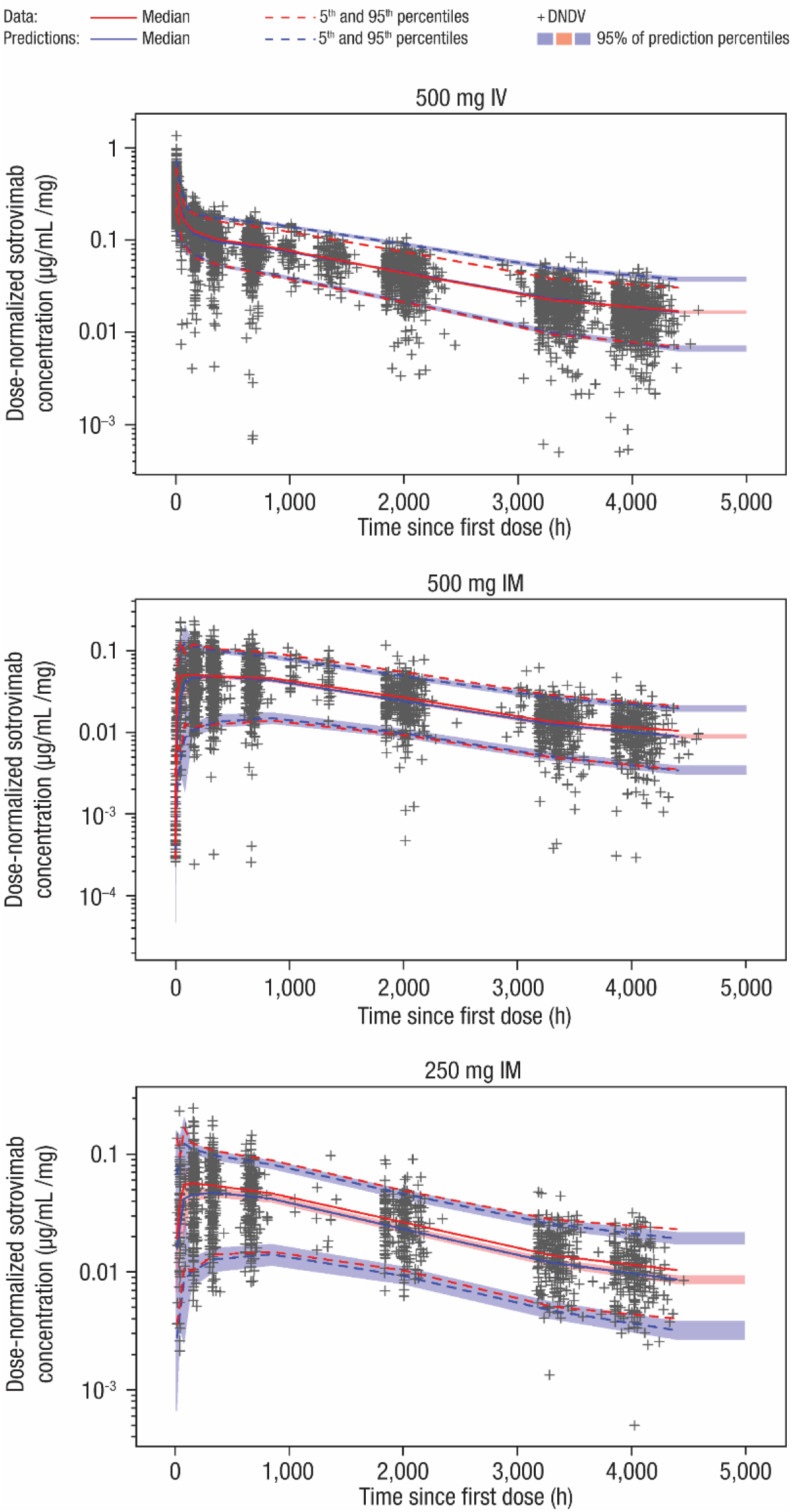
Visual predictive check plots for the final population PK model of sotrovimab. CI, confidence interval; PK, pharmacokinetic.

The magnitude of covariate effects on key exposure measures (C_max_, C_96h_, C_168h_) were summarized using forest plots (**Figure 3 and Figures S2 and S3**). For all significant covariates (body weight, BMI, sex), the GMR (90% CI) ranged between 0.63-1.21 (0.58-1.34); thus, were fully contained within prespecified relevance bounds of 0.5 to 2.0. Additionally, for all other evaluated covariates, nearly all GMRs and associated 90% CIs fell within the 0.5 to 2.0 bounds, with only a few exceptions. The upper bound of the 90% CI exceeded 2.0 for the IM exposures C_96h_ and C_168h_ for participants who self-identified as Asian, however, the GMR and 90% CI for C_max_ were <2.0. The lower bound of the 90% CI was below 0.5 in the IM dose group for two other covariate subgroups: participants with a race classification of “other” and participants who received remdesivir or dexamethasone. However, the numbers of participants in these subgroups were limited and are not expected to allow reliable estimation of the relevance of these covariates on exposure. Overall, the majority of covariates investigated were contained within the pre-specified 0.5 to 2.0 bounds.

**FIGURE 3.**
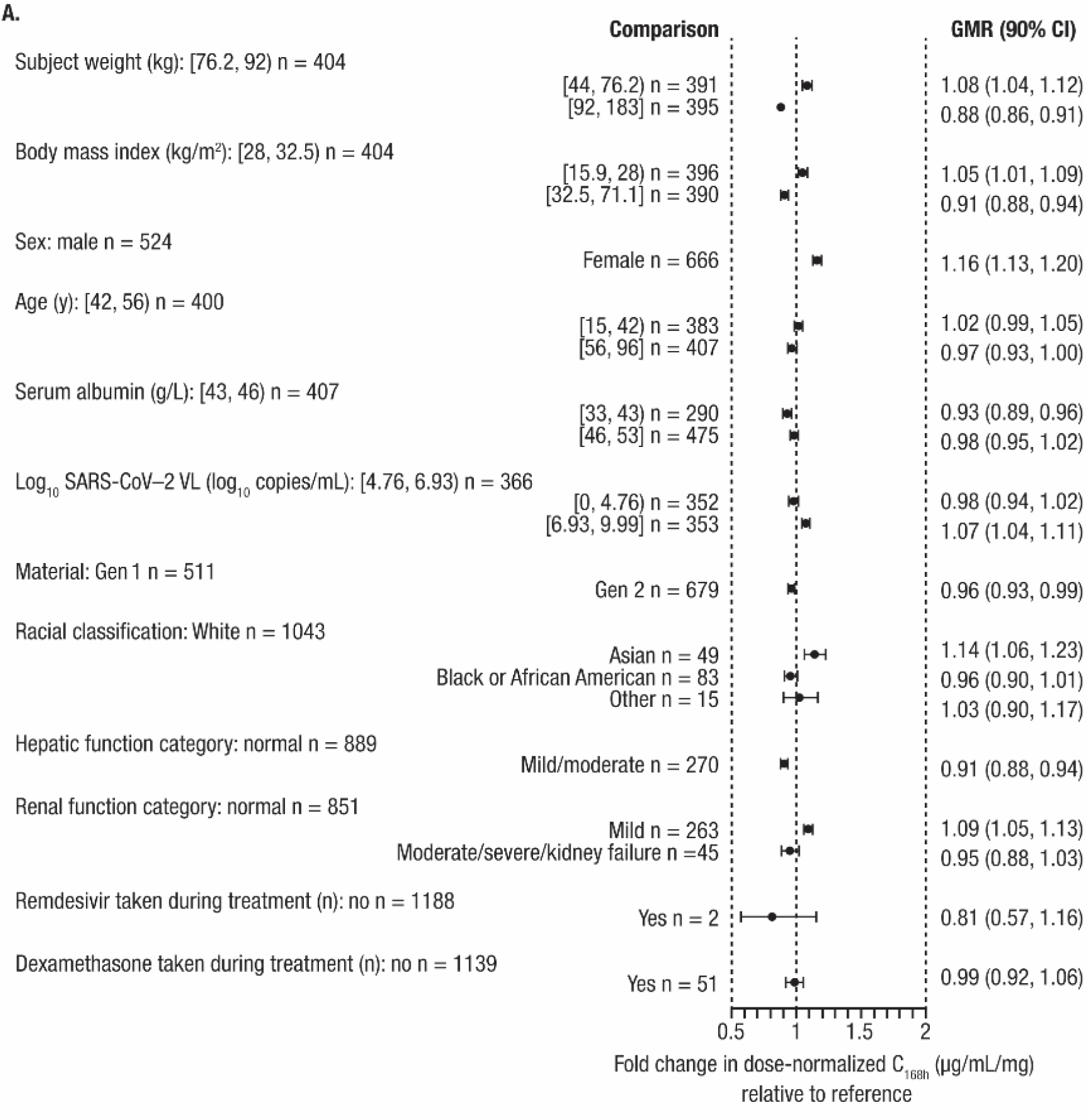
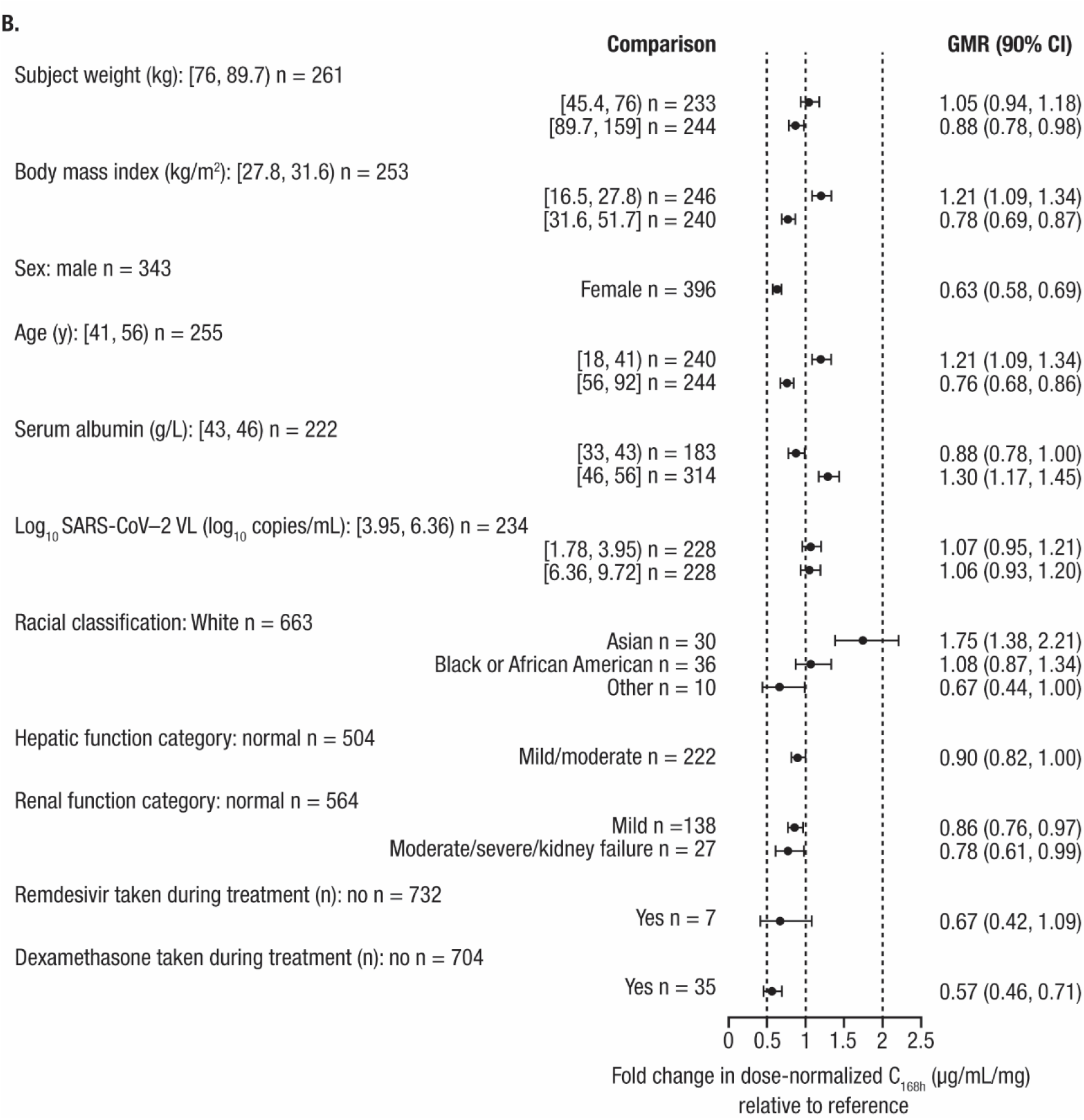
Forest plots of geometric mean ratios (90% confidence interval) of model-estimated dose-normalized C_0-168h_, after (A) IV and (B) IM dosing. n is the number of patients in each group, [ or ] indicates respective endpoint is included in the interval, and (or) indicates respective endpoint is not included in the interval. C_168h_, concentration at 168 hours; CI, confidence interval; GMR, geometric mean ratio; IM, intramuscular; SARS-CoV-2, severe acute respiratory syndrome coronavirus 2; VL, viral load.

### ER modeling

The source data for the ER analyses included 2,877 records of 959 patients from COMET-TAIL. After excluding patients who were not in the intent-to-treat population and removing patients lacking PK data, a total of 2,706 observations from 902 patients were included in the COMET-TAIL ER efficacy dataset for these analyses. **Table S6** summarizes the numbers of patients included in the efficacy analyses, by treatment group.

Summaries of demographic characteristics for the ER efficacy analysis population are provided in **Table S7**. Among the 902 patients, 493 (45.3%) were male, median age was 50 years, and median baseline BMI was 31.0 kg/m^2^. The median duration of symptoms was 4 days, and median log_10_ baseline SARS-CoV-2 viral load was 6.09 (range, 3.2 to 10.2). An estimated 69.8% of patients had ≤1 risk factor and 30.2% had >1 risk factor. Overall, 2.2% of patients had progression of COVID-19 through Day 29 (**Table 2**), with the highest percentage in the sotrovimab 250 mg IM arm (4.0%).

**TABLE 2.** Summary statistics of progression occurrence rates for the primary endpoint in the COMET-TAIL study, by dose (ER population)

| Endpoint |  | Sotrovimab<br>(250 mg IM) | Sotrovimab<br>(500 mg IM) | Sotrovimab<br>(500 mg IV) | Overall |
| --- | --- | --- | --- | --- | --- |
| Progression<br>of COVID-19, n (%) | No | 167 (96.0) | 353 (97.8) | 362 (98.6) | 882 (97.8) |
|  | Yes | 7 (4.0) | 8 (2.2) | 5 (1.4) | 20 (2.2) |

COVID-19, coronavirus disease 2019; ER, exposure-response; IM, intramuscular, IV, intravenous.

The exposure measures of sotrovimab AUC_0□Day28_, C_96h,_ and C_168h_ were significant predictors of the probability of progression (*p* < 0.05). The linear functions of sotrovimab C_96h_ and C_168h_ were selected for further model development, as both were considered clinically meaningful exposure parameters based on timing of progression (median time from randomization to progression event was 5.5 days). Parameter estimates and standard errors from the base ER progression of COVID-19 through Day 29 models of sotrovimab C_168h_ and C_96h_ are shown in **Table S8**.

For each treatment arm, the base model was used to predict the progression rates associated with the respective range of exposures. **Table S9** shows that for the C_168h_ model, within each treatment arm the observed progression rate was encompassed by the range of predicted progression rates. Conversely, for the C_96h_ model, while the observed progression rate in the ER dataset fell within the predicted range for the 500 mg IV and 500 mg IM treatment arms, predicted progression rates fell below the observed point estimate in the 250 mg IM treatment arm (**Table S10**). Therefore, the C_168h_ model will be the focus of the remainder of the ER results summary.

Covariate analysis led to the addition of the number of risk factors (≤1 vs >1) as an additive shift on the model intercept (model-estimated placebo response). The impact of number of risk factors was only on the intercept (placebo progression rate), with no impact on drug response.

Parameter estimates and standard errors from the final ER model (progression of COVID-19 through Day 29 model vs sotrovimab C_168h_) are shown in **Table 3**. All parameters were estimated with good precision (<55 %RSE).

**TABLE 3.** Parameter estimates and standard errors from the final ER model for the occurrence of progression of COVID-19 through Day 29 (primary endpoint) – sotrovimab concentrations at 168 hours in the COMET-TAIL study

| Parameter |  | Final parameter estimate |  |
| --- | --- | --- | --- |
| 168 hours |  | Population mean | %RSE |
| INT | Overall response (logit) (-) | -4.169 | 12.03 |
|  | Additive shift in INT for<br>RISKCATN = 1 | 1.887 | 27.34 |
| SLP | Slope for concentration at 168<br>hours (1/[µg/mL]) | -0.02037 | 54.43 |
| Minimum value of the objective function = 170.913 |  |  |  |

COVID-19, coronavirus disease 2019; ER, exposure-response; RISKCATN, number of risk factors; %RSE, relative standard error expressed as a percentage.

Using the ER efficacy analysis dataset in conjunction with the parameter estimates from the final C_168h_ model, 500 replicates of the analysis dataset were simulated. **Figure 4** illustrates the predicted probability of progression of COVID-19 through Day 29 (primary endpoint) and 95% CI from the simulated datasets (blue line) overlaid on the observed proportion of patients with progression (red line) versus sotrovimab C_168h_. The observed proportion of data fell within the 95% CI of predicted proportions across the range of sotrovimab concentrations, indicating an adequate model fit.

**FIGURE 4.**
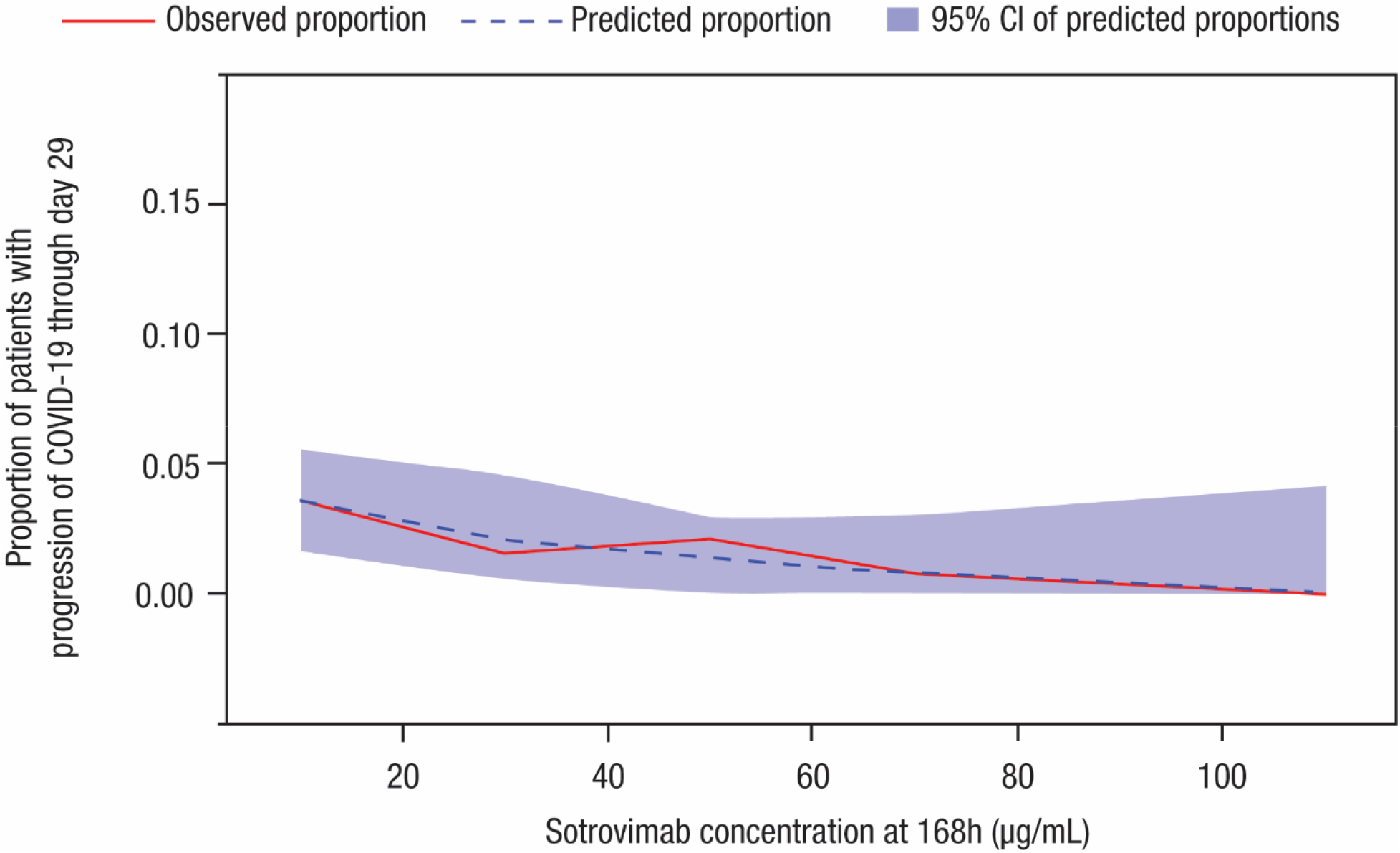
Visual predictive check plots for the final exposure-response model for the occurrence of COVID-19 progression through Day 29 (primary endpoint) versus sotrovimab concentrations at 168 hours. CI, confidence interval; COVID-19, coronavirus disease 2019.

## DISCUSSION

Understanding the factors influencing both exposure and response of therapeutic agents can inform dosing recommendations when considered alongside cumulative safety and efficacy data. A key objective of this analysis was to establish a popPK model for sotrovimab to identify sources of variability in exposures. Additionally, individual exposure measures for patients in COMET-TAIL were predicted using the popPK model and included in the ER analysis dataset to establish an ER relationship for the primary efficacy endpoint (probability of progression of COVID-19) and identify any potential sources of variability in drug response.

Sotrovimab PK was described by a linear, two-compartment model with first-order elimination, and IM absorption was characterized by a sigmoid absorption model. A systematic covariate analysis found that body weight was a statistically significant descriptor of the variability in sotrovimab PK and influenced the CL and V3. Other covariate effects retained in the final model were sex on KA, and F_IM_ and BMI on KA. However, the magnitudes of covariate effects are not expected to be clinically relevant based on available data to date. Additional safety and efficacy data could inform the clinical relevance of covariate effect on exposure.

The ER analyses aimed to assess the relationship between sotrovimab serum exposure and clinical response of progression of COVID-19, as defined as hospitalization >24 hours or death. ER models for the probability of progression of COVID-19 through Day 29 (primary endpoint) using only data from COMET-TAIL were developed. VPC plots showed that the observed proportion of data fell within the 95% CI of predicted proportions across the range of sotrovimab concentrations at 168 hours, indicating an adequate model fit. Furthermore, the number of risk factors (≤1 vs >1) as an additive shift on intercept was the only significant covariate effect. This covariate impacted only the model-estimated placebo response (intercept) but had no impact on overall drug response.

However, despite the COMET-TAIL model performance, the ER analysis has a number of limitations that may prevent the generalization of these results to describe the overall exposure-progression relationship for sotrovimab in early treatment across SARS-COV-2 variants. The COMET-TAIL models included a limited number of progressors (20 progressors, 882 non-progressors). Notably, PK data were not collected in several progressors (3/10 in 250 mg IM arm, 2/10 in 500 mg IM arm); therefore, the ER dataset is smaller than the efficacy dataset, resulting in differences in progression rates/treatment arm between the clinical efficacy and ER datasets. The COMET-TAIL models allowed for assessment of ER for only one predominant variant of concern (VOC). COMET-TAIL recruitment was from June to September 2021, when the predominant circulating strain of SARS-CoV-2 in the United States was the delta variant.^21^ Consistent with variant circulation at the time of study enrollment, the predominant VOC/variant of interest detected in COMET-TAIL participants with available sequencing data was the delta (B.1.617.2) variant (88.2%, 674/764 participants) (data on file). The translatability of ER analyses from delta variant to other variants is unknown. Additionally, given the difference in early PK between IV and IM, an AUC-based exposure measure (AUC_0-168h_) would be expected to more appropriately capture the difference in exposure between IV and IM. However, with the exception of AUC_Day0-28_, which was not considered to be clinically relevant due to timing of progressions (median time from randomization to event was 5.5 days), the exposure measures identified as statistically significant predictors of response were single concentration exposure measures (C_168h_ and C_96h_). As these timepoints represent a period where IV and IM PK profiles are converging and do not account for higher early exposures following IV administration, it is expected that the ER model may provide a conservative estimate of target IV exposures needed to achieve the desired efficacy. Furthermore, COMET-TAIL lacked a placebo arm; therefore, the model intercept was informed by limited data available at low exposures. The model-predicted placebo progression rate was also lower than expected considering the largely unvaccinated high-risk COMET-TAIL study population who were predominantly recruited in the state of Florida, United States (85%) from June to September 2021, when the predominant circulating strain of SARS-CoV-2 in the United States was the delta variant.^21^ Real-world evidence (RWE) data point to an estimated placebo progression rate of 9.1% in unvaccinated individuals in Florida during the delta period (data on file).

Therefore, while the ER analysis provided some initial insights into exposure measures influencing response and factors expected to influence placebo progression rates in COMET-TAIL, the dataset may be too limited to directly inform the exposure-progression relationship for sotrovimab in early treatment and dosing recommendations for current or future VOCs. Not only does the limited efficacy dataset limit the model’s utility, but the translatability of exposure response analysis conducted in the context of a single variant to future VOCs requires further investigation across the field. Therefore, the consideration of cumulative evidence from in vitro neutralization, clinical safety, RWE of effectiveness, and popPK data may be needed to guide future dosing recommendations.

## STUDY HIGHLIGHTS

### What is the current knowledge on the topic?

The efficacy of sotrovimab in preventing the progression of COVID-19 in non-hospitalized patients with mild to moderate COVID-19 at high risk for disease progression has been evaluated in two pivotal clinical trials.

### What question did this study address?

What are the sources of variability in sotrovimab exposure and exposure-response (ER)? What is the relationship between sotrovimab serum exposure and prevention of progression of COVID-19?

### What does this study add to our knowledge?

Body weight, sex, and BMI were covariates of sotrovimab exposure but are not anticipated to be clinically relevant. Sotrovimab concentrations at 96 and 168 hours are significant predictors of COVID-19 progression. Number of risk factors is a covariate of the ER model-predicted placebo progression rate, with no impact on drug effect.

### How might this change drug discovery, development, and/or therapeutics?

The results of these analyses inform on sources of variability in sotrovimab exposure as well as the exposure measures found to be predictive of progression of mild-moderate COVID-19 in early treatment of COVID-19.

## Data Availability

Individual participant data will not be made publicly available. The authors confirm that the data supporting the findings of this study are available within the article and its supplementary materials.

## Acknowledgements

The authors thank Michelle Preston, MSc, and Jeanne McKeon, PhD, of Lumanity Scientific, Inc., for medical writing support, which was funded by Vir Biotechnology, Inc., and GSK.

## Author Contributions

JES, AE-Z, JP, SR, MA, AN, AS, EA, WWY, EM, CG, AP, AES, and MR designed the research. JES, AE-Z, JP, and SR performed the research. JES, AE-Z, JP, SR, and XL analyzed the data. All authors wrote the manuscript.

## Footnotes

a A post hoc change was made to the multiple imputation algorithm from daily to weekly imputation due to the bias that was observed in the imputed progression rates from the daily imputation algorithm.

