## Supplemental Materials for "Population pharmacokinetics and exposure-response analysis of sotrovimab in the early treatment of COVID-19"

**SUPPLEMENTARY INFORMATION TITLES**

**Derived sotrovimab pharmacokinetic (PK) parameters based on the final model**

**TABLE S1** PK sampling during the included clinical studies

**TABLE S2** Planned evaluation of covariates in analyses

**TABLE S3** Participants and sotrovimab concentration data included in the population PK model

**TABLE S4** Demographics and disease characteristics of the PK population

**TABLE S5** Summary statistics of derived PK parameters

**TABLE S6** Data disposition for exposure-response COMET-TAIL efficacy dataset

**TABLE S7** Summary statistics of demographic characteristics in the COMET-TAIL study

**TABLE S8** Parameter estimates and standard errors from the base exposure-response model for the occurrence of progression of COVID-19 through Day 29 (primary endpoint) – sotrovimab concentrations at 168 and 96 hours in the COMET-TAIL study

**TABLE S9** Summary statistics of the model-predicted probability of COVID-19 progression through Day 29 (primary endpoint) versus treatment for the base sotrovimab concentrations at 168 hours exposure-response model (COMET-TAIL)

**TABLE S10** Summary statistics of the model-predicted probability of COVID-19 progression through Day 29 (primary endpoint) versus treatment for the base sotrovimab concentrations at 96 hours exposure-response model (COMET-TAIL)

**FIGURE S1** Goodness-of-fit plots for the final population PK model of sotrovimab, stratified by (A) IV and (B) IM route of administration

**FIGURE S2** Forest plots of geometric mean ratios (90% CI) of model-estimated dose-normalized C_96h_, after (A) IV and (B) IM dosing

**FIGURE S3** Forest plots of geometric mean ratios (90% CI) of model-estimated dose-normalized C_max_, after (A) IV and (B) IM dosing

**MODEL CODE S1** Final Population PK Model of Sotrovimab

**MODEL CODE S2** Final ER Model of Sotrovimab 168 hours in the COMET-TAIL study

**MODEL CODE S3** Final ER Model of Sotrovimab 96 hours in the COMET-TAIL study

**Derived sotrovimab pharmacokinetic (PK) parameters based on the final model**

Key PK parameters steady-state volume of distribution (V_SS_) and terminal half-life (t_½_) were calculated.

The V_SS_ was calculated as the sum of the central and peripheral volumes of distribution. The t_½_ was calculated using the micro-rate constants as described in Toutain et al.^1^

**TABLE S1** PK sampling during the included clinical studies

| Study | Study population | Sotrovimab dosing | PK sampling plan |
| --- | --- | --- | --- |
| COMET-ICE | Patients with mild to moderate COVID-19 | 500 mg IV | Dense sampling for 169 days (n = 10)  Sparse sampling (n = 660) |
| COMET-TAIL | Patients with mild to moderate COVID-19 | 500 mg IV, 250 mg IM, 500 mg IM | Sparse sampling up to 169 days |
| COMET-PEAK | Patients with mild to moderate COVID-19 | 500 mg IV, 250 mg IM, 500 mg IM | Dense sampling for lead-in cohort; sparse sampling for expansion cohort (up to 169 days) |
| BLAZE-4 | Patients with mild to moderate COVID-19 | 500 mg IV | Sparse sampling up to 85 days |
| GSK Study 217653 | Healthy Japanese and White participants | 500 mg IV, 500 mg IM | Dense sampling up to 127 days |

COVID-19, coronavirus disease 2019; IM, intramuscular; IV, intravenous; PK, pharmacokinetic.

**TABLE S2** Planned evaluation of covariates in analyses

| Covariate | Pharmacokinetic parameters | | |
| --- | --- | --- | --- |
|  | Absorption, bioavailability | Clearance | Volume of distribution |
| Age | X | X | X |
| Body weight | X | X | X |
| BMI | X | X | X |
| NCI hepatic function category | X | X | X |
| Renal function category | X | X | X |
| Sex | X | X | X |
| Sotrovimab clinical trial material (Gen 1 or Gen 2)^a^ | X | X | X |
| Serum albumin | X | X | X |
| Race | X | X | X |
| Disease state (COVID-19/healthy) | X | X | X |
| Concomitant medication | X | X | X |
| Baseline viral load | X | X | X |

^a^Gen 1 was a clinical drug substance that was generated from a stable pool of cells. Gen 2 is the commercial drug substance generated from a single clone from Gen 1.

BMI, body mass index; COVID-19, coronavirus disease 2019; NCI, National Cancer Institute.

**TABLE S3** Participants and sotrovimab concentration data included in the population PK model

| Study | Study population | Sotrovimab dosing | Total number of participants | Sotrovimab concentration data points included |
| --- | --- | --- | --- | --- |
| COMET-ICE | Patients with mild to moderate COVID-19 | 500 mg IV | 503 | 3,127 |
| COMET-TAIL | Patients with mild to moderate COVID-19 | 500 mg IV | 383 | 2,054 |
|  |  | 250 mg IM | 185 | 1,003 |
|  |  | 500 mg IM | 377 | 2,011 |
| COMET-PEAK | Patients with mild to moderate COVID-19 | 500 mg IV | 191 | 1,644 |
|  |  | 250 mg IM | 76 | 531 |
|  |  | 500 mg IM | 81 | 571 |
| BLAZE-4 | Patients with mild to moderate COVID-19 | 500 mg IV | 95 | 312 |
| GSK Study 217653 | Healthy Japanese and White participants | 500 mg IV | 18 | 251 |
|  |  | 500 mg IM | 20 | 256 |

IM, intramuscular; IV, intravenous; PK, pharmacokinetic.

**TABLE S4** Demographics and disease characteristics of the PK population

| Characteristic | Participants (N = 1,929) |
| --- | --- |
| COVID-19 patients, n (%) | 1,891 (98.0) |
| Mean (SD) log_10_ SARS-CoV-2 viral load, log_10_ copies/mL | 4.96 (2.58) |
| Healthy participants, n (%) | 38 (2.0) |
| Sex, n (%) |  |
| Male | 867 (44.9) |
| Female | 1,062 (55.1) |
| Age, years |  |
| Median (range) | 49.0 (15, 96) |
| Self-reported race, n (%) |  |
| White | 1,706 (88.4) |
| Black/African American | 119 (6.2) |
| Asian | 79 (4.1) |
| Other | 13 (0.7) |
| Median (range) body weight, kg | 83.6 (44.0, 183.0) |
| Median (range) body mass index, kg/m^2^ | 30.4 (15.9, 71.1) |
| NCI hepatic function category, n (%) |  |
| Normal | 1,393 (72.2) |
| Mild | 487 (25.2) |
| Moderate | 5 (0.3) |
| Missing | 44 (2.3) |
| BSA normalized renal function category, n (%) |  |
| Normal | 1,415 (73.4) |
| Mild | 401 (20.8) |
| Moderate | 65 (3.4) |
| Severe | 5 (0.3) |
| Renal failure | 2 (0.1) |
| Missing | 41 (2.1) |
| Mean (SD) serum albumin, g/L | 44.77 (3.40) |
| Concomitant dexamethasone, n (%) | 86 (4.5) |
| Concomitant remdesivir, n (%) | 9 (0.5) |
| Sotrovimab treatment, n (%) |  |
| 500 mg IV | 1,190 (61.7) |
| 500 mg IM | 478 (24.8) |
| 250 mg IM | 261 (13.5) |
| Sotrovimab clinical trial material^a^, n (%) |  |
| Gen 1 | 511 (26.5) |
| Gen 2 | 1,418 (73.5) |

^a^Gen 1 was a clinical drug substance that was generated from a stable pool of cells. Gen 2 is the commercial drug substance generated from a single clone from Gen 1.

BSA, body surface area; COVID-19, coronavirus disease 2019; IM, intramuscular; IV, intravenous; NCI, National Cancer Institute; PK, pharmacokinetic; SARS-CoV-2, severe acute respiratory syndrome coronavirus 2; SD, standard deviation.

**TABLE S5** Summary statistics of derived PK parameters

| Exposures | Overall |
| --- | --- |
| Clearance (L/day) | N = 1,927 |
| Mean (SD) | 0.101 (0.050) |
| Geometric mean (geometric %CV) | 0.095 (34.996) |
| Median | 0.091 |
| 5^th^, 95^th^ | 0.06, 0.17 |
| Central volume of distribution (L) | N = 1,927 |
| Mean (SD) | 3.786 (5.679) |
| Geometric mean (geometric %CV) | 3.287 (43.887) |
| Median | 3.177 |
| 5^th^, 95^th^ | 1.89, 6.26 |
| Peripheral volume of distribution (L) | N = 1,927 |
| Mean (SD) | 4.616 (1.540) |
| Geometric mean (geometric %CV) | 4.429 (28.285) |
| Median | 4.411 |
| 5^th^, 95^th^ | 2.87, 6.89 |
| Steady-state volume of distribution (L) | N = 1,927 |
| Mean (SD) | 8.402 (6.131) |
| Geometric mean (geometric %CV) | 7.881 (30.700) |
| Median | 7.696 |
| 5^th^, 95^th^ | 5.32, 12.53 |
| Terminal half-life (d) | N = 1,927 |
| Mean (SD) | 61.410 (9.837) |
| Geometric mean (geometric %CV) | 60.670 (15.746) |
| Median | 61.236 |
| 5^th^, 95^th^ | 47.84, 75.12 |
| IM bioavailability fraction | N = 739 |
| Mean (SD) | 0.476 (0.174) |
| Geometric mean (geometric %CV) | 0.435 (49.995) |
| Median | 0.497 |
| 5^th^, 95^th^ | 0.18, 0.72 |

%CV, coefficient of variation expressed as a percent; IM, intramuscular; N, number of virtual patients; PK, pharmacokinetic; SD, standard deviation.

**TABLE S6** Data disposition for ER COMET-TAIL efficacy dataset

| Reason for exclusion | Records excluded | Patients affected | Patients excluded | Number following exclusion | |
| --- | --- | --- | --- | --- | --- |
|  |  |  |  | Records | Patients |
| Source efficacy data |  |  |  | 2,877 | 959 |
| Remove patients not in ITT population | 87 | 29 | 29 | 2,790 | 930 |
| Remove patients with no exposure measures | 84 | 28 | 28 | 2,706 | 902 |
| Total |  |  |  | 2,706 | 902 |

ER, exposure-response; ITT, intent-to-treat.

**TABLE S7** Summary statistics of demographic characteristics in the COMET-TAIL study

| Patient characteristic | N = 902 |
| --- | --- |
| Age, years |  |
| Mean (SD) | 50.1 (16.7) |
| Median (min, max) | 50.0 (15, 92) |
| Males, n (%) | 409 (45.3) |
| Body mass index at baseline, kg/m^2^ |  |
| Mean (SD) | 30.8 (5.2) |
| Median (min, max) | 31.0 (18, 63) |
| Duration of symptoms, days |  |
| Mean (SD) | 3.6 (1.6) |
| Median (min, max) | 4.0 (0, 7) |
| Symptom duration category, n (%) |  |
| ≤3 days | 442 (49.0) |
| 4-5 days | 344 (38.1) |
| >5 days | 116 (12.9) |
| Log_10_ baseline SARS-CoV-2 viral load |  |
| Mean (SD) | 5.90 (1.98) |
| Median (min, max) | 6.09 (3.2, 10.2) |
| Number of risk factors, grouped, n (%) |  |
| ≤1 | 630 (69.8) |
| >1 | 272 (30.2) |
| Number of other risk factors, n (%) |  |
| 0 | 604 (67.0) |
| 1 | 265 (29.4) |
| 2 | 29 (3.2) |
| 3 | 4 (0.4) |
| Route of administration, n (%) |  |
| IV | 367 (40.7) |
| IM | 535 (59.3) |

IM, intramuscular, IV, intravenous; SARS-CoV-2, severe acute respiratory syndrome coronavirus 2; SD, standard deviation.

**TABLE S8** Parameter estimates and standard errors from the base ER model for the occurrence of progression of COVID-19 through Day 29 (primary endpoint) – sotrovimab concentrations at 168 and 96 hours in the COMET-TAIL study

| Parameter |  | Final parameter estimate | |
| --- | --- | --- | --- |
| 168 hours |  | Population mean | %RSE |
| INT | Overall response (logit) (-) | -3.03 | 12.1 |
| SLP | Slope for concentration at 168 hours (1/[μg/mL]) | -0.0255 | 46.4 |
| Minimum value of the objective function = 186.391 | | | |
| 96 hours |  | Population mean | %RSE |
| INT | Overall response (logit) (-) | -3.187 | 10.41 |
| SLP | Slope for concentration at 96 hours (1/[μg/mL]) | -0.01838 | 50.54 |
| Minimum value of the objective function = 187.084 | | | |

COVID-19, coronavirus disease 2019; ER, exposure-response; %RSE, relative standard error expressed as a percentage.

**TABLE S9** Summary statistics of the model-predicted probability of COVID-19 progression through Day 29 (primary endpoint) versus treatment for the base sotrovimab concentrations at 168 hours ER model (COMET-TAIL)

| Model-predicted probability of progression | Sotrovimab  250 mg IM (N = 174) | Sotrovimab  500 mg IM (N = 361) | Sotrovimab  500 mg IV (N = 367) | Overall  (N = 902) |
| --- | --- | --- | --- | --- |
| Mean (SD) | 0.0340 (0.0066) | 0.0267 (0.0089) | 0.0120 (0.0054) | 0.0221 (0.0113) |
| Median (min, max) | 0.0337  (0.013, 0.045) | 0.0266  (0.003, 0.046) | 0.0111  (0.002, 0.044) | 0.0204  (0.002, 0.046) |
| 10^th^, 90^th^ percentile | 0.026, 0.042 | 0.015, 0.039 | 0.007, 0.017 | 0.009, 0.039 |

COVID-19, coronavirus disease 2019; ER, exposure-response; IM, intramuscular; IV, intravenous; SD, standard deviation.

**TABLE S10** Summary statistics of the model-predicted probability of COVID-19 progression through Day 29 (primary endpoint) versus treatment for the base sotrovimab concentrations at 96 hours ER model (COMET-TAIL)

| Model-predicted probability of progression | Sotrovimab  250 mg IM (N=174) | Sotrovimab  500 mg IM (N=361) | Sotrovimab 500 mg IV  (N=367) | Overall  (N=902) |
| --- | --- | --- | --- | --- |
| Mean (SD) | 0.0325 (0.0047) | 0.0278 (0.0067) | 0.0117 (0.0048) | 0.0222 (0.0104) |
| Median (min, max) | 0.0329  (0.015, 0.039) | 0.0284  (0.005, 0.039) | 0.0107  (0.003, 0.038) | 0.0228  (0.003, 0.039) |
| 10^th^, 90^th^ percentile | 0.027, 0.038 | 0.018, 0.036 | 0.008, 0.017 | 0.009, 0.036 |

ER, exposure-response; IM, intramuscular; IV, intravenous; SD, standard deviation.

**FIGURE S1** Goodness-of-fit plots for the final population pharmacokinetic model of sotrovimab, stratified by (A) IV and (B) IM route of administration. IM, intramuscular; IV, intravenous; |IWRES|, absolute value of the individual weighted residuals; NPDE, normalized prediction distribution error.

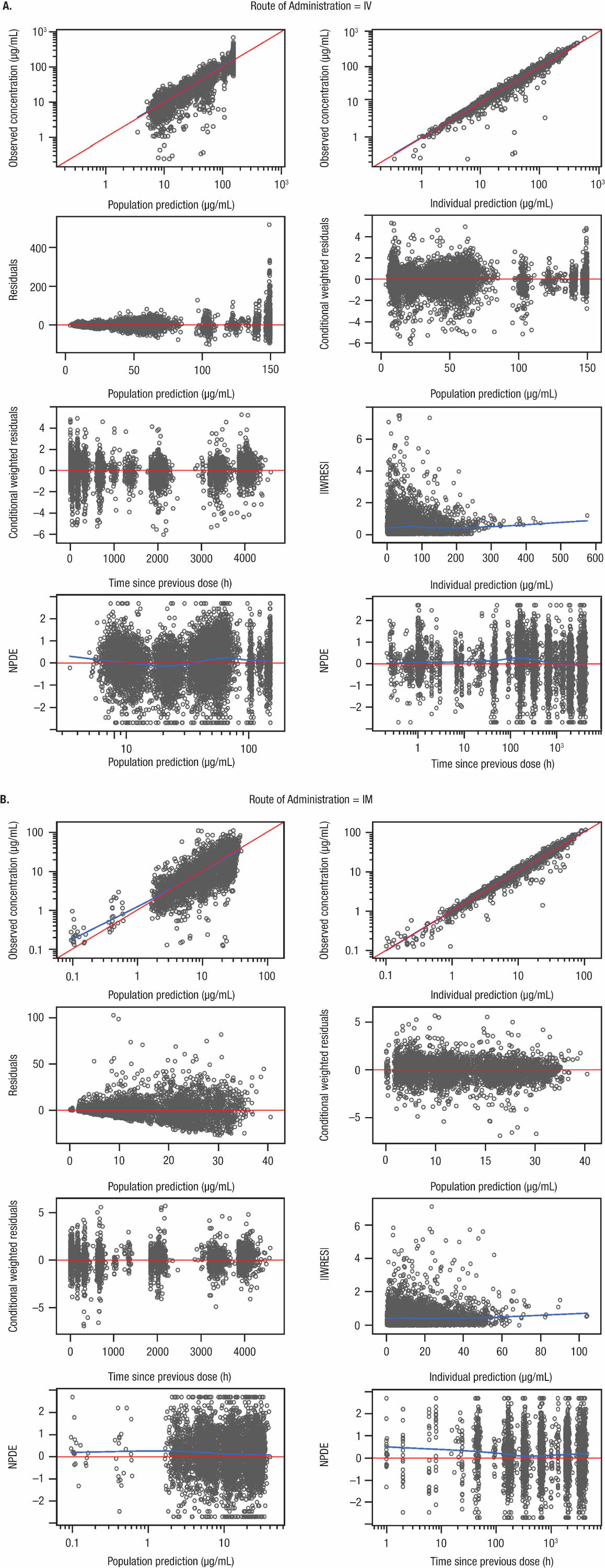

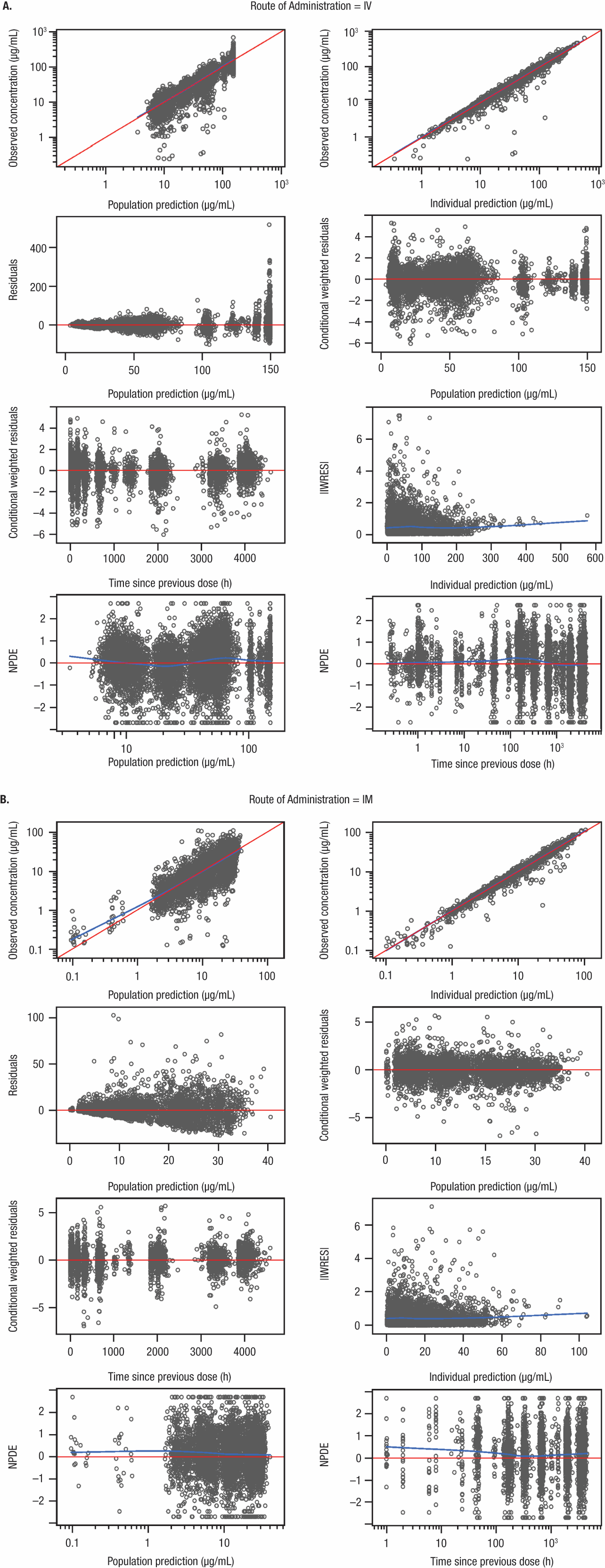

**FIGURE S2** Forest plots of geometric mean ratios (90% CI) of model-estimated dose-normalized C_96h_, after (A) IV and (B) IM dosing.

n is the number of patients in each group, [ or ] indicates respective endpoint is included in the interval, and ( or ) indicates respective endpoint is not included in the interval.

C_96h_, concentration at 96 hours; CI, confidence interval; GMR, geometric mean ratio; IM, intramuscular; IV, intravenous; SARS-CoV-2, severe acute respiratory syndrome coronavirus 2; VL, viral load.

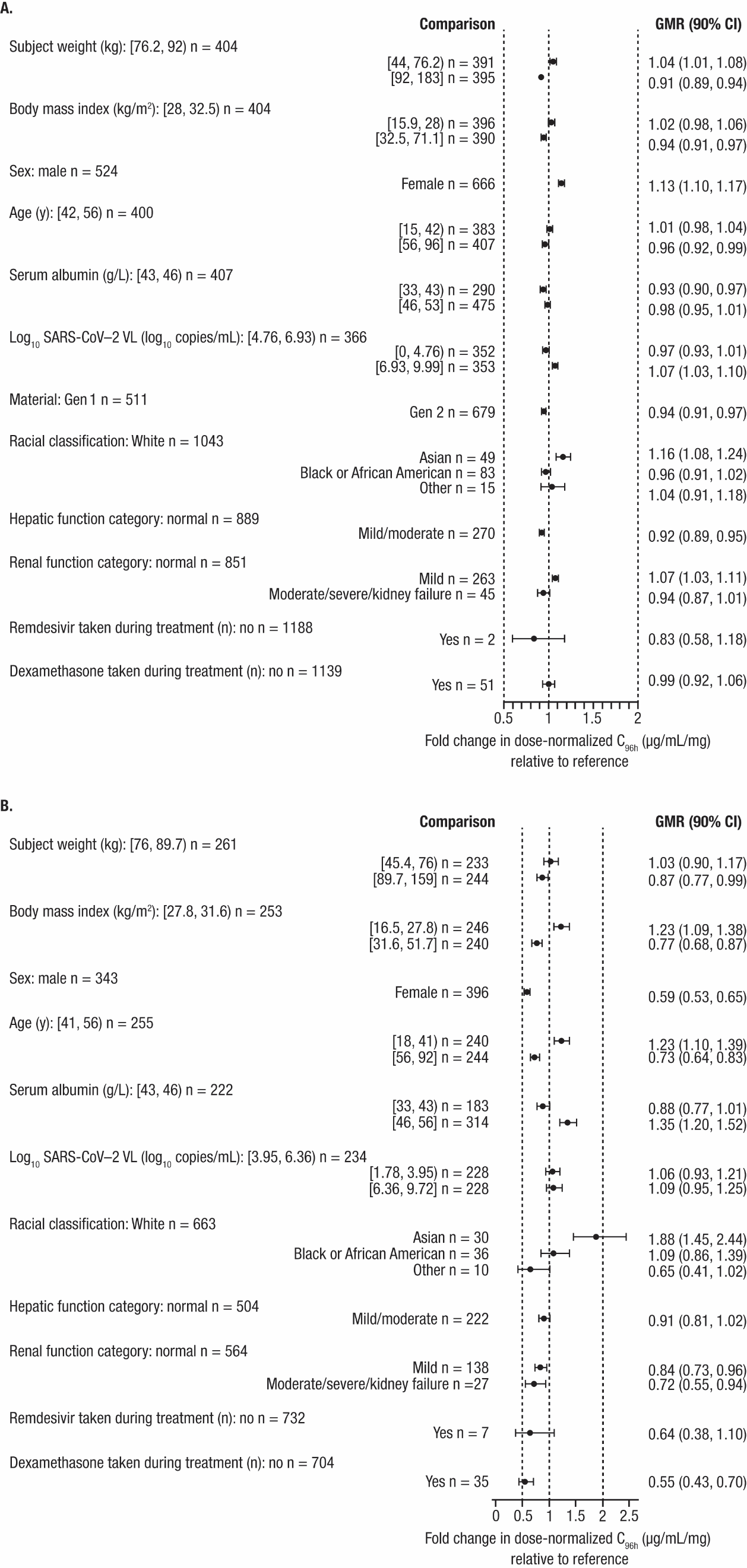

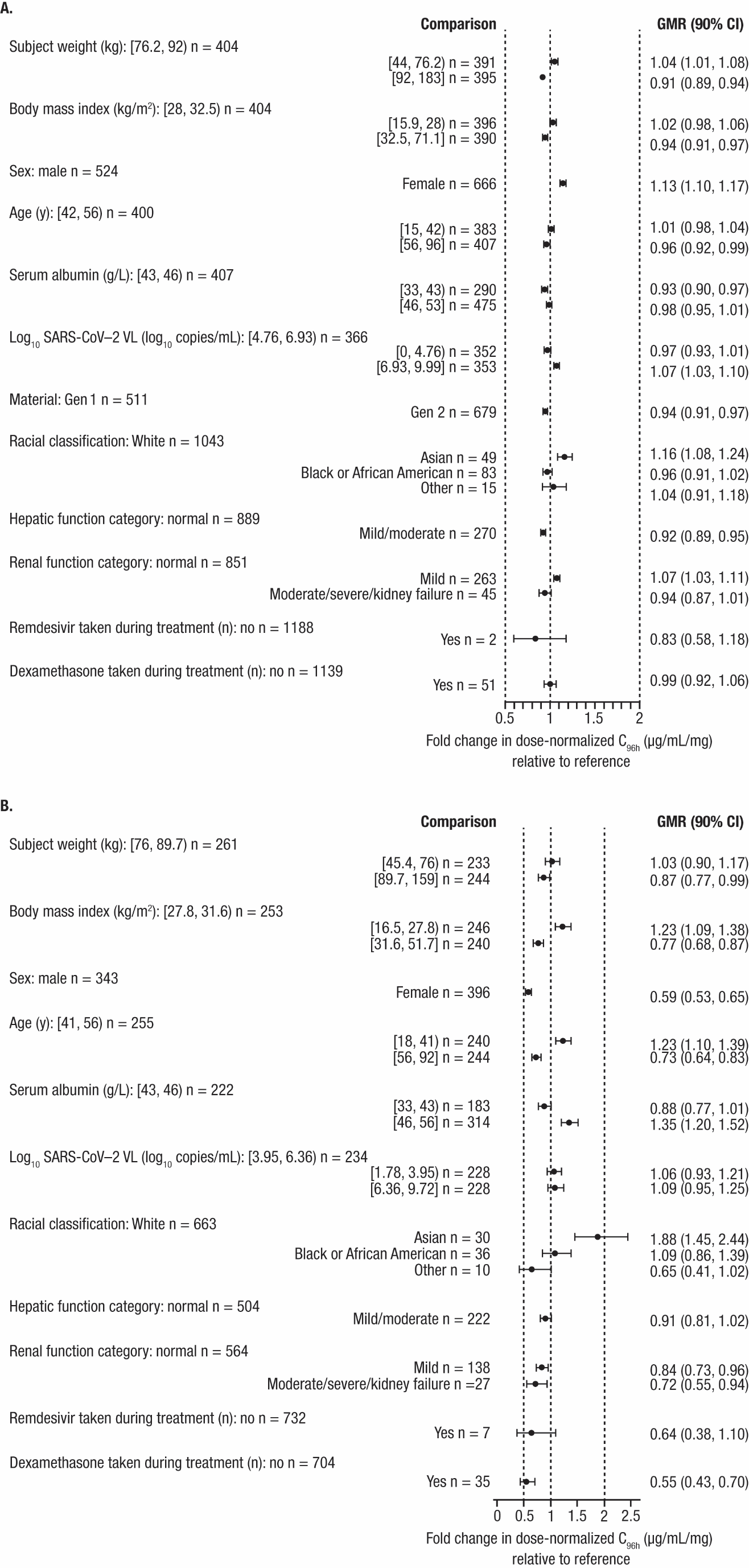

**FIGURE S3** Forest plots of geometric mean ratios (90% confidence interval) of model-estimated dose-normalized C_max_, after (A) IV; and (B) IM dosing.

n is the number of patients in each group, [ or ] indicates respective endpoint is included in the interval, and ( or ) indicates respective endpoint is not included in the interval.

CI, confidence interval; C_max_, maximum concentration; GMR, geometric mean ratio; IM, intramuscular; IV, intravenous; SARS-CoV-2, severe acute respiratory syndrome coronavirus 2; VL, viral load.

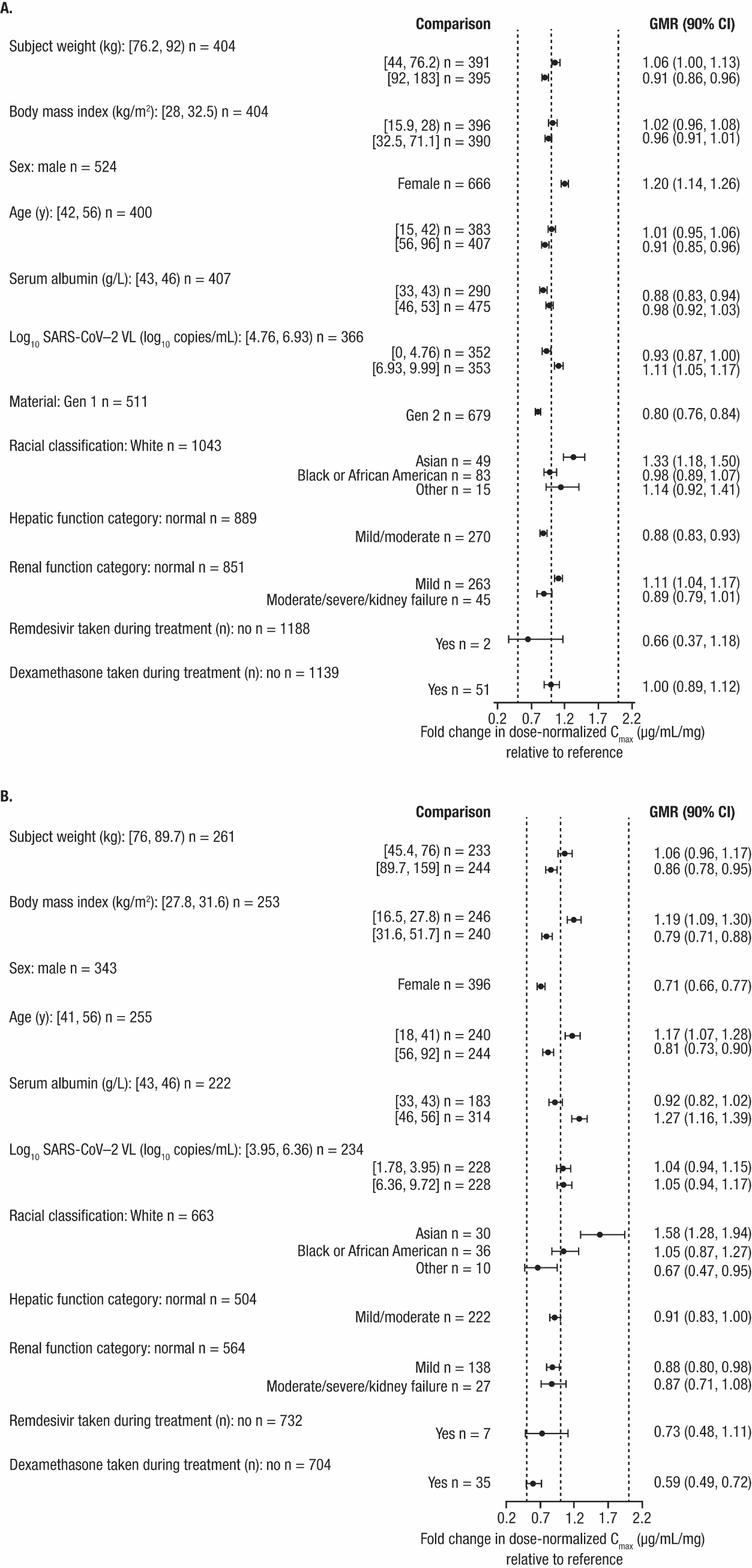

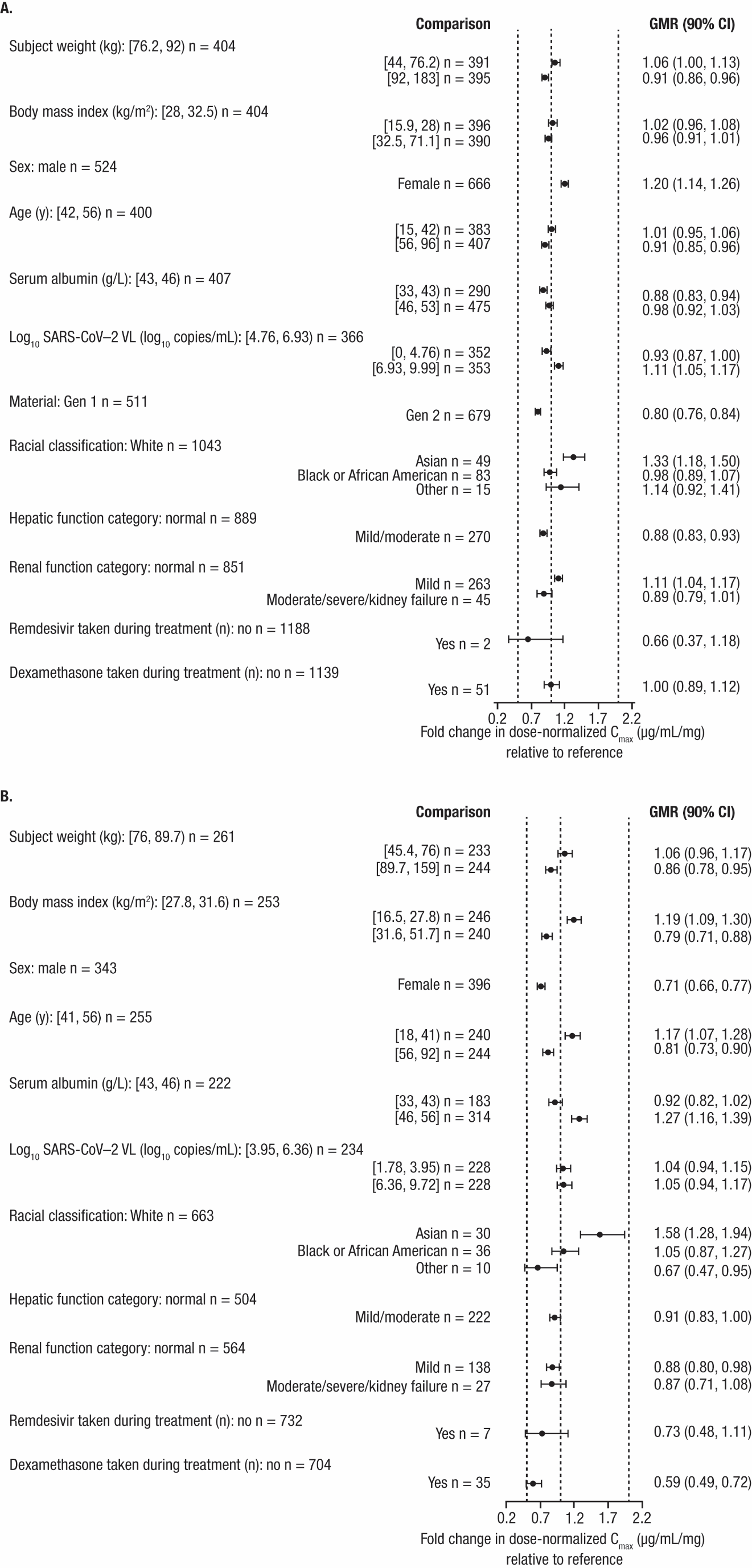

**MODEL CODE S1** Final Population PK Model of Sotrovimab

$PROBLEM fin005 ;; Written for use with NONMEM Version 7

;; Covariate effects

;; 1: IWTBL on V3 as POWER

;; 2: IWTBL on CL as POWER

;; 3: IIBMIBL on KA as POWER

;; 4: SEXF on LBIOAV as ADD

;; 5: SEXF on KA as PROP

;;

;; PURPOSE: model development

;; Sotrovimab administered IV, IM

;; LLOQ: 100 mcg/mL

;; -----------------------------------------------------------------------------

$INPUT C ID TIME DV LNDV DVID AMT RATE1 RATE ROUTE MAT TRTM DUR CMT MDV EVID NOMT VISN BLQ DOSE ADA SEQN AGEBL SEXN IARACEN FDARACEN ETHN IWTBL IHTBL IIBMIBL IBSADUBL IBMIGRN2 IEGFRBBL ITPROTBL IALBBL RENIMPBN HEPIMPCN VLCOVBL REMFLN DEXFLN PKFN NOEPK NOEPK2 SPARSE DENSE ACTARM PLBFLN STUDYN SUBJIDN USUBJID=DROP DAT=DROP TIME=DROP

$DATA data.csv

IGNORE=@

IGNORE=(BLQ.EQ.1)

IGNORE=(PKFN.EQ.0)	;Placebo subjects

IGNORE=(CMT.EQ.4)

IGNORE=(SEQN.EQ.5257, SEQN.EQ.5258)

IGNORE=(SEQN.EQ.85, SEQN.EQ.102, SEQN.EQ.1302, SEQN.EQ.2426, SEQN.EQ.2807, SEQN.EQ.2823, SEQN.EQ.3212, SEQN.EQ.3983)

IGNORE=(SEQN.EQ.3984, SEQN.EQ.4220, SEQN.EQ.5084, SEQN.EQ.5343, SEQN.EQ.5541, SEQN.EQ.6398, SEQN.EQ.6664, SEQN.EQ.6681)

IGNORE=(SEQN.EQ.6691, SEQN.EQ.6774, SEQN.EQ.7026, SEQN.EQ.7313, SEQN.EQ.7890, SEQN.EQ.8463, SEQN.EQ.10083)

IGNORE=(SEQN.EQ.10765, SEQN.EQ.11268, SEQN.EQ.13147, SEQN.EQ.13399, SEQN.EQ.13539, SEQN.EQ.13555, SEQN.EQ.13557)

IGNORE=(SEQN.EQ.13563, SEQN.EQ.13571, SEQN.EQ.13579, SEQN.EQ.13587, SEQN.EQ.13736, SEQN.EQ.14480, SEQN.EQ.15273)

IGNORE=(SEQN.EQ.15652, SEQN.EQ.16123, SEQN.EQ.16217)

IGNORE=(SEQN.EQ.2830, SEQN.EQ.3985, SEQN.EQ.5050, SEQN.EQ.6524, SEQN.EQ.6655, SEQN.EQ.6673, SEQN.EQ.6798, SEQN.EQ.6807)

IGNORE=(SEQN.EQ.7226, SEQN.EQ.7786, SEQN.EQ.8261, SEQN.EQ.8728, SEQN.EQ.11255, SEQN.EQ.11266, SEQN.EQ.13570, SEQN.EQ.13572)

IGNORE=(SEQN.EQ.13903, SEQN.EQ.14481, SEQN.EQ.15646, SEQN.EQ.16218)

IGNORE=(SEQN.EQ.1993, SEQN.EQ.3986, SEQN.EQ.6637, SEQN.EQ.6776, SEQN.EQ.7189, SEQN.EQ.7190, SEQN.EQ.10267, SEQN.EQ.12015)

IGNORE=(SEQN.EQ.13540, SEQN.EQ.13547, SEQN.EQ.13580, SEQN.EQ.13904, SEQN.EQ.16122, SEQN.EQ.7191, SEQN.EQ.10268, SEQN.EQ.16121)

IGNORE=(ID.EQ.197, ID.EQ.235, ID.EQ.530, ID.EQ.535, ID.EQ.536, ID.EQ.538, ID.EQ.543, ID.EQ.544, ID.EQ.545, ID.EQ.547)

IGNORE=(ID.EQ.558, ID.EQ.562, ID.EQ.564, ID.EQ.733, ID.EQ.734, ID.EQ.735, ID.EQ.875, ID.EQ.897, ID.EQ.1148, ID.EQ.1279)

IGNORE=(ID.EQ.1281, ID.EQ.1378, ID.EQ.1424, ID.EQ.1524, ID.EQ.1555, ID.EQ.1582, ID.EQ.1650, ID.EQ.1729, ID.EQ.1739, ID.EQ.1849)

IGNORE=(ID.EQ.1885, ID.EQ.1945, ID.EQ.1986, ID.EQ.1987, ID.EQ.2028, ID.EQ.2030, ID.EQ.2133, ID.EQ.2134, ID.EQ.2136, ID.EQ.2140)

IGNORE=(ID.EQ.2162, ID.EQ.2189, ID.EQ.2246, ID.EQ.2287, ID.EQ.2316, ID.EQ.2317, ID.EQ.2336, ID.EQ.2348, ID.EQ.2367, ID.EQ.2368)

IGNORE=(ID.EQ.2544, ID.EQ.2546, ID.EQ.2564, ID.EQ.2570, ID.EQ.2605)

$SUBROUTINES ADVAN6 TRANS1 TOL=9

$MODEL

COMP=(depot)

COMP=(central, DEFOBS)

COMP=(periph)

$THETA

(0, 0.096)	;--th1- CL: Elimination Clearance in Participants of 83.6 kg (L/day)

(0, 3.5)	;--th2- V2: Central Volume of Distribution (L)

(0, 0.65)	;--th3- Q: Distribution Clearance (L/day)

(0, 4.5)	;--th4- V3: Peripheral Volume of Distribution in Participants of 83.6 kg (L)

(0, 0.0057)	;--th5- KA: Absorption Rate in Male Participants with BMI of 30.41 m^2 (1/h)

(0, 0.5)	;--th6- FIM: Intra-Muscular Bioavailability in Male Participants

(0, 150)	;--th7- R1: Rate of Input (mg/h)

(-INF, 0.76)	;--th8- V3: Power of Body Weight Effect

(-INF, 0.58)	;--th9- CL: Power of Body Weight Effect

(-INF, -1.1)	;--th10- KA: Power of BMI Effect

(-INF, -0.1)	;--th11- FIM: Additive Shift in Female Participants

(-1, 0.01)	;--th12- KA: Proportional Shift in Female Participants

$OMEGA BLOCK(5)

0.1	;--eta1- IIV in CL [expp]

0.01 0.3	;--eta2- IIV in V2 [expp]

0.01 0.01 0.9	;--eta3- IIV in FIM [cv=100*(1-th6)*eta3]

0.01 0.01 0.01 0.3	;--eta4- IIV in KA [expp]

0.01 0.01 0.01 0.01 0.1	;--eta5- IIV in V3 [expp]

$SIGMA

0.2	;--eps1- Constant CV RV component [accv1=eps1-ccv;eps2-add]

0.5	;--eps2- Additive RV component	[accv1]

$PK

STUDY=67 ;ICE

IF (STUDYN.EQ.216912) STUDY=12 ;PEAK

IF (STUDYN.EQ.217079) STUDY=79 ;BLAZE

IF (STUDYN.EQ.217114) STUDY=14 ;TAIL

IF (STUDYN.EQ.217653) STUDY=53 ;Japan

;Sex, male (SEXN==1) is reference

SEXF = 1

IF (SEXN.EQ.1) SEXF=0

ALB = IALBBL ;(g/L)

IF (IALBBL.LE.0) ALB=45

MRACE = 4; other

IF (FDARACEN.EQ.2) MRACE=1 ;Asian

IF (FDARACEN.EQ.3) MRACE=2 ;Black

IF (FDARACEN.EQ.5) MRACE=3 ;White

;Disease state HEALTHY=0 is reference

HEALTHY = 0

IF (STUDYN.EQ.217653) HEALTHY=1

;Redefine viral load

VLBL = VLCOVBL

IF (VLCOVBL.LT.0) VLBL = 5.276 ;set to median if missing in patients

IF (VLCOVBL.EQ.0) VLBL = 1.78 ;set to minimum if 0 in patients

IF (STUDYN.EQ.217653) VLBL = 1.78 ;healthy

IMFLN = 0 ;IM flag

IF (ROUTE.EQ.2) IMFLN = 1

;-------------------------------------------------------

;--COVARIATE EFFECTS

COV5 = (1+THETA(12)*SEXF)

COV4 = THETA(11)*SEXF

COV3 = (IIBMIBL/30.41)**THETA(10)

COV2 = (IWTBL/83.6)**THETA(9)

COV1 = (IWTBL/83.6)**THETA(8)

TVCL = THETA(1) * COV2

CL = TVCL * EXP(ETA(1)) ;(L/day)

TVV2 = THETA(2)

V2 = TVV2 * EXP(ETA(2))

TVQ = THETA(3)

Q = TVQ	;(L/day)

TVV3 = THETA(4) * COV1

V3 = TVV3 * EXP(ETA(5))

TVKA = THETA(5) * COV3 * COV5

KA = 0 ;IV

IF (ROUTE.EQ.2) KA = TVKA * EXP(ETA(4))

TVFIM = THETA(6)

TVLFIM = log (TVFIM / (1-TVFIM)) + COV4

FIM=1

IF (ROUTE.EQ.2) THEN

LFIM = TVLFIM + ETA(3)

FIM = EXP(LFIM) / (1 + EXP(LFIM)) ;IM bioavailability

END IF

F1 = FIM

TVR1 = THETA(7)

R1=0

IF (ROUTE.EQ.2) R1 = TVR1

R2=0

IF (ROUTE.EQ.1) R2 = RATE

;;; Exclude the etas forced to 0 from the eta shrinkage calculations ;;;

IF (ROUTE==1) ETASXI(3)=1 ;exclude IV data from shrinkage calculation of IIV in LBIOAV

IF (ROUTE==2) ETASXI(3)=2 ;include all IM data in the shrinkage calculation of IIV in LBIOAV

IF (ROUTE==1) ETASXI(4)=1 ;exclude IV data from shrinkage calculation of IIV in KA

IF (ROUTE==2) ETASXI(4)=2 ;include all IM data in the shrinkage calculation of IIV in KA

K20 = (CL/24)/V2

K23 = (Q/24)/V2

K32 = (Q/24)/V3

; SCALING FACTOR

; AMT = mg

; VOL = L

; CONC = mcg/mL

; SCALE = V2

S2 = V2

DNDV = DV / DOSE

;Derived parameters

AUC = DOSE / CL ;mcg*day/mL

IF (ROUTE.EQ.2) AUC = F1 * DOSE / CL

VSS = V2 + V3

CLH = CL / 24 ;CL in L/h

;Calculate half-life

TERM1 = K32 + K23 + K20

TERM2 = 4 * K32 * K20

NUMER = TERM1 - SQRT (TERM1**2 - TERM2)

BETA = NUMER/2

HALF = (LOG(2)/BETA) / 24 ;in days

STRT=0 ;should not occur

IF (ROUTE.EQ.1 .AND. DOSE.EQ.500) STRT=1

IF (ROUTE.EQ.2 .AND. DOSE.EQ.250) STRT=2

IF (ROUTE.EQ.2 .AND. DOSE.EQ.500) STRT=3

STRTA = 1000*STRT + STUDY

$DES

DADT(1) = -KA * A(1)

DADT(2) = KA * A(1) -K20 * A(2) -K23 * A(2) +K32 * A(3)

DADT(3) = K23 * A(2) -K32 * A(3)

$ERROR

IPRED = F

IRES = DV - IPRED

W = SQRT(IPRED**2*SIGMA(1,1) + SIGMA(2,2))

IWRES = IRES/W

Y = IPRED + IPRED*EPS(1) + EPS(2)

$ESTIMATION METHOD=CONDITIONAL INTERACTION PRINT=1 MAXEVAL=9999 NSIG=3 SIGL=9 SORT MSFO=fin005.msf

$COVARIANCE MATRIX=S PRINT=E UNCONDITIONAL ;MATRIX=R Fisher information matrix

$TABLE ID SUBJIDN STUDY ACTARM TIME NOMT DNDV EVID MDV SEQN VISN DOSE ROUTE TRTM DUR TVCL TVV2 TVQ TVV3 TVKA TVFIM TVLFIM TVR1

CL V2 Q V3 KA LFIM FIM F1 R1 ETA1 ETA2 ETA3 ETA4 ETA5 MAT ADA MRACE HEALTHY STRT STRTA AUC VSS CLH HALF

AGEBL SEXN IARACEN FDARACEN ETHN IWTBL IHTBL IIBMIBL IBSADUBL IBMIGRN2 IEGFRBBL ITPROTBL IALBBL

ALB RENIMPBN HEPIMPCN REMFLN DEXFLN VLCOVBL VLBL

IPRED IRES IWRES CWRES NPDE ONEHEADER NOPRINT FILE=fin005.tbl FORMAT=s1PE12.6

**MODEL CODE S2** Final ER Model of Sotrovimab 168 hours in the COMET-TAIL study

```
$PROBLEM tail-primary-eff-base-model-cp168

;; PURPOSE: Logistic regression model for Primary Eff - Progression of COVID-19
;; one record per patient
;; Study COMET-TAIL only
;; Base Model - Linear Function of Cp168
;; -----------------------------------------------------------------------------

$INPUT STUDY NUM ID MDV EVID PDE DV AVAL ACTIVE DOSE GTRT ADY DSFD AGE SEXF
BMIBL DROP=IMPBMIBL RFNUMCND RISKCATN RFBMI30N RFAGE55N NOTHRISK TTEVENT
SYMDUR SYMDCTN VLCOVBL DROP=IMPVLBL VARIANTN ROUTEN BNEUT BLYMPH BNEUTLYM
AUC024 AUC048 AUC072 AUC096 AUC0168 AUC0D28 CP24 CP48 CP72 CP96 CP168
DROP=CAVG24 DROP=CAVG48 DROP=CAVG72 DROP=CAVG96 DROP=CAVG168 DROP=AVISITN
REGION1N REGION2N AAGE DROP=AGEGR1N DROP=AGEGR2N DROP=AGEGR3N DROP=AGEGR4N
DROP=AGEGR5N DROP=AGEGR6N SEXN ASEXN DROP=RACEGR1N DROP=RACEGR2N ARACEN
DROP=TRT01PN DROP=TRT01AN DROP=TRTPN DROP=TRTAN DROP=FUDAYS DROP=BMIBLCTN
DROP=BMIBLC2N DROP=VLCVBCTN DROP=VLCVBC2N DROP=RFDIABFN DROP=RFOBESFN
DROP=RFCKDFN DROP=RFCHFFN DROP=RFNCGR1N DROP=RFNCGR2N DROP=HEPIMPN
DROP=RENIMPN DROP=FPSYMN DROP=SEROPBLN DROP=SUBJIDN DROP=MSID DROP=USUBJID


$DATA ../../data/ereff.csv
IGNORE=@
IGNORE=(PDE.NE.1)  ;Keep PDE=1 only
IGNORE=(STUDY.EQ.214367) ;Keep COMET-TAIL only 217114

$THETA
-3.5        ;--th1- INT: Overall Response (logit) (-)
-0.02       ;--th2- SLP: Slope for CP168 (1/[ug/mL])

$OMEGA
0 FIX  ;--eta1- IIV in INT [add]


;--parms- INT SLP
;--randparms- ETA1
;--contcovs- AGE BMIBL SYMDUR VLCOVBL TTEVENT
;--catcovs- SEXF RISKCATN NOTHRISK SYMDCTN VARIANTN ROUTEN


$PRED
  ; PD parameters
  TVINT = THETA(1)
  INT = TVINT

  ;DRUG effect
  TVSLP= THETA(2)
  SLP = TVSLP
  DRUG=SLP*CP168

  ; Logit of cumulative probabilities
  LOGIT = INT + DRUG + ETA(1)

  ; Cumulative probabilities
  PROB = EXP(LOGIT)/(1 + EXP(LOGIT))

  ; Likelihood estimation
  Y = PROB
  IF (DV == 0) Y = 1-PROB

$ESTIMATION METHOD=COND LIKELIHOOD LAPLACIAN PRINT=1 MAXEVAL=9999 NSIG=3 SORT

$COVARIANCE PRINT=E

$TABLE STUDY NUM ID MDV EVID PDE AVAL ACTIVE DOSE GTRT ADY DSFD AGE SEXF
BMIBL RFNUMCND RISKCATN RFBMI30N RFAGE55N NOTHRISK TTEVENT SYMDUR SYMDCTN
VLCOVBL VARIANTN ROUTEN BNEUT BLYMPH BNEUTLYM AUC024 AUC048 AUC072 AUC096
AUC0168 AUC0D28 CP24 CP48 CP72 CP96 CP168 ARACEN PROB TVINT INT
ETA1 TVSLP SLP DRUG ONEHEADER NOPRINT FILE=tail-primary-eff-base-model-cp168.tbl
```

**MODEL CODE S3** Final ER Model of Sotrovimab 96 hours in the COMET-TAIL study

```
$PROBLEM tail-primary-eff-base-model-cp96

;; PURPOSE: Logistic regression model for Primary Eff - Progression of COVID-19
;; one record per patient
;; Study COMET-TAIL only
;; Base Model - Linear Function of Cp96
;; -----------------------------------------------------------------------------

$INPUT STUDY NUM ID MDV EVID PDE DV AVAL ACTIVE DOSE GTRT ADY DSFD AGE SEXF
BMIBL DROP=IMPBMIBL RFNUMCND RISKCATN RFBMI30N RFAGE55N NOTHRISK TTEVENT
SYMDUR SYMDCTN VLCOVBL DROP=IMPVLBL VARIANTN ROUTEN BNEUT BLYMPH BNEUTLYM
AUC024 AUC048 AUC072 AUC096 AUC0168 AUC0D28 CP24 CP48 CP72 CP96 CP168
DROP=CAVG24 DROP=CAVG48 DROP=CAVG72 DROP=CAVG96 DROP=CAVG168 DROP=AVISITN
REGION1N REGION2N AAGE DROP=AGEGR1N DROP=AGEGR2N DROP=AGEGR3N DROP=AGEGR4N
DROP=AGEGR5N DROP=AGEGR6N SEXN ASEXN DROP=RACEGR1N DROP=RACEGR2N ARACEN
DROP=TRT01PN DROP=TRT01AN DROP=TRTPN DROP=TRTAN DROP=FUDAYS DROP=BMIBLCTN
DROP=BMIBLC2N DROP=VLCVBCTN DROP=VLCVBC2N DROP=RFDIABFN DROP=RFOBESFN
DROP=RFCKDFN DROP=RFCHFFN DROP=RFNCGR1N DROP=RFNCGR2N DROP=HEPIMPN
DROP=RENIMPN DROP=FPSYMN DROP=SEROPBLN DROP=SUBJIDN DROP=MSID DROP=USUBJID


$DATA ../../data/ereff.csv
IGNORE=@
IGNORE=(PDE.NE.1)  ;Keep PDE=1 only
IGNORE=(STUDY.EQ.214367) ;Keep COMET-TAIL only 217114

$THETA
-3.5        ;--th1- INT: Overall Response (logit) (-)
-0.02       ;--th2- SLP: Slope for CP96 (1/[ug/mL])

$OMEGA
0 FIX  ;--eta1- IIV in INT [add]


;--parms- INT SLP
;--randparms- ETA1
;--contcovs- AGE BMIBL SYMDUR VLCOVBL TTEVENT
;--catcovs- SEXF RISKCATN NOTHRISK SYMDCTN VARIANTN ROUTEN


$PRED
  ; PD parameters
  TVINT = THETA(1)
  INT = TVINT

  ;DRUG effect
  TVSLP= THETA(2)
  SLP = TVSLP
  DRUG=SLP*CP96

  ; Logit of cumulative probabilities
  LOGIT = INT + DRUG + ETA(1)

  ; Cumulative probabilities
  PROB = EXP(LOGIT)/(1 + EXP(LOGIT))

  ; Likelihood estimation
  Y = PROB
  IF (DV == 0) Y = 1-PROB

$ESTIMATION METHOD=COND LIKELIHOOD LAPLACIAN PRINT=1 MAXEVAL=9999 NSIG=3 SORT

$COVARIANCE PRINT=E

$TABLE STUDY NUM ID MDV EVID PDE AVAL ACTIVE DOSE GTRT ADY DSFD AGE SEXF
BMIBL RFNUMCND RISKCATN RFBMI30N RFAGE55N NOTHRISK TTEVENT SYMDUR SYMDCTN
VLCOVBL VARIANTN ROUTEN BNEUT BLYMPH BNEUTLYM AUC024 AUC048 AUC072 AUC096
AUC0168 AUC0D28 CP24 CP48 CP72 CP96 CP168 ARACEN PROB TVINT INT
ETA1 TVSLP SLP DRUG ONEHEADER NOPRINT FILE=tail-primary-eff-base-model-cp96.tbl
```
